# Inherited human TFIIIA deficiency disrupts T cell development

**DOI:** 10.64898/2026.07.01.26356670

**Authors:** Evi Duthoo, Sueun Park, Tamara Jarayseh, Marita Bosticardo, Rafah Mackeh, Yana Van Droogenbroeck, Imke Velghe, Veronique Debacker, Hailun Li, Nourhen Agrebi, Francesca Pala, Sarah Ghistelinck, Julie Braet, Sandra Van Lint, Lukas Pieters, Martin Watelet, Hajar Besbassi, Leslie Naesens, Tessa Kerre, Delfien Bogaert, Hye Sun Kuehn, Sergio D. Rosenzweig, Emmanuelle Jouanguy, Ottavia M. Delmonte, Ivan K. Chinn, Stephen Hughes, Amel Hassan, Mohammed Y. Karim, Asha Elmi, Giuliana Giardino, Claudio Pignata, Bénédicte Neven, Benson Ogunjimi, Karim Vermaelen, Denis L.J. Lafontaine, Mirjam van der Burg, Anne Puel, Jérémie Rosain, Jean-Laurent Casanova, Bernice Lo, Patrick Sips, Tom Taghon, Jacinta Bustamante, Luigi D. Notarangelo, Simon J. Tavernier, Filomeen Haerynck

## Abstract

Molecular characterization of human monogenic inborn errors of T cell immunity provides both biological insights and medical progress. We report rare biallelic deleterious variants in *GTF3A*, encoding transcription factor IIIA (TFIIIA), a zinc-finger protein required for transcription and chaperoning of 5S ribosomal RNA (rRNA). These variants were identified in ten patients from eight unrelated families and eight countries presenting with either T^-^B^+^NK^+^ severe combined immunodeficiency (SCID) or combined immune deficiency (CID), characterized by T cell lymphopenia and variable antibody deficiency. The *GTF3A* variants disrupt TFIIIA function through distinct mechanisms, including defective DNA binding, aberrant nuclear localization, and reduced protein stability compromising TFIIIA-mediated transcription and chaperoning of 5S rRNA. Using artificial thymic organoids derived from TFIIIA-deficient CD34⁺ progenitors, an early developmental arrest at the T cell commitment stage was documented *in vitro*. Zebrafish deficient for *gtf3aa* recapitulated the impaired thymocyte development *in vivo*. Together, these findings establish TFIIIA deficiency as a novel cause of (S)CID, expanding the genetic and mechanistic landscape of inborn errors of T cell immunity and uncovering an essential role for TFIIIA in human adaptive immunity.

**One-sentence summary:** Biallelic *GTF3A* variants causing TFIIIA deficiency, which disrupts 5S rRNA transcription, represent a novel monogenic cause of (S)CID, unveiling a crucial role of TFIIIA in T cell development.

## INTRODUCTION

The study of inborn errors of immunity (IEIs) has been transformative in uncovering genes and pathways indispensable for human immune defense and homeostasis (*1*, *2*). Predominantly caused by single-gene defects, IEIs illustrate how disruption of a single pathway can perturb human immunity, leading to a wide spectrum of clinical phenotypes (*3*). Severe combined immunodeficiency (SCID) and combined immunodeficiency (CID) represent the most severe IEIs and are characterized by impaired T cell development and/or function, often accompanied by variable impairment of the B and NK cell compartments. SCID typically presents during early infancy with profound susceptibility to life threatening infections. CID manifests later in life with a more heterogeneous clinical course, in which infections are often compounded by immune dysregulation, including autoimmunity, autoinflammation, allergy, and lymphoproliferation (*3–5*). Without early recognition and targeted treatment, these conditions are often fatal (*6*, *7*).

Despite advances in our understanding of the molecular underpinning of (S)CID, 30-40% of affected patients still lack a definitive molecular diagnosis (*3*, *8*). Accurate molecular diagnosis is essential for guiding early supportive care and curative treatment including hematopoietic stem cell transplantation (HSCT), gene therapy, enzyme replacement therapy, or thymus transplantation (*8*, *9*). Beyond its clinical implications, this persistent diagnostic gap highlights an incomplete understanding of the molecular pathways governing T cell development and function. Identifying novel genetic causes of (S)CID is therefore essential, not only to improve diagnostic precision and inform targeted therapeutic approaches, but also to advance fundamental insight into adaptive immune biology, particularly T cell development.

Transcription factor IIIA (TFIIIA), encoded by *GTF3A*, is uniquely responsible for transcription of 5S ribosomal RNA (rRNA), a core structural component of the 60S large ribosomal subunit (*10*). This ∼42 kDa C_2_H_2_-type zinc-finger protein contains nine tandem zinc-finger motifs that mediate both DNA and RNA binding (*11–13*). TFIIIA binds the internal control region (ICR) of 5S ribosomal DNA (rDNA) genes and subsequently recruits TFIIIB and TFIIIC to initiate transcription of 5S rRNA by RNA polymerase III (*11*, *14*). Beyond its transcriptional role, TFIIIA acts as a chaperone, stabilizing newly synthesized 5S rRNA and directing it to sites of ribosome assembly (*15–17*). While decades of work in model organisms have clarified many aspects of TFIIIA biology (*12*, *16*, *18–20*), whether and how disruption of this ubiquitously required process leads to human disease has remained largely unexplored.

In 2022, we reported two siblings from the first family with biallelic variants (p.C195W/p.C219R) in *GTF3A*, presenting with an IEI characterized by increased susceptibility to herpes simplex encephalitis (HSE) (*21*). Mechanistic studies demonstrated that TFIIIA transcriptionally regulates *RNA5SP141*, a 5S rRNA pseudogene that acts as an endogenous ligand for the cytosolic RNA sensor retinoic acid-inducible gene-I (RIG-I), which is involved in type I interferon (IFN) production upon viral infection (*21*, *22*). These findings revealed a previously unrecognized role for TFIIIA in innate antiviral defense. Notably, both affected siblings reported in the study also exhibited antibody deficiency and B and T cell maturation defects, raising the possibility that TFIIIA function extends beyond innate immune signaling and may contribute more broadly to lymphocyte biology.

Here, we identify TFIIIA deficiency as a novel monogenic cause of (S)CID, uncovering a hitherto unrecognized role for TFIIIA in T cell ontogeny.

## RESULTS

### Personally identifiable patient information was redacted in accordance with medRxiv requirements

#### Identification of biallelic *GTF3A* variants in patients presenting with (S)CID

We identified a cohort of ten patients from eight unrelated families presenting with clinical and immunological features of SCID or CID, characterized by recurrent or invasive infections and T cell lymphopenia (Fig. 1A, Tables 1-3). All four SCID patients (P1-P4) presented during infancy, with three of four diagnosed through newborn screening. The six CID patients (P5-10) presented during childhood. Across the cohort, all patients developed severe viral, bacterial, and/or fungal infections predominantly affecting the respiratory and/or gastrointestinal tracts (detailed information on identified pathogens is provided in Table 2). Of note, herpes simplex virus type 1 (HSV-1)-associated disease was observed in two related (siblings) CID patients, manifesting as HSV-1 stomatitis (P7) and HSV-1 encephalitis (P8) (*21*). One adolescent (P5) developed features of immune dysregulation, including ulcerative colitis and disseminated skin granuloma positive for rubella (secondary to MMR vaccination) and human papillomavirus (HPV). No consistent non-immune features indicative of syndromic disorder or skeletal dysplasia were observed, although P3 presented with intrauterine growth restriction (IUGR) and P5 presented with short stature and delayed neurocognitive development, likely secondary to chronic illness and neonatal hypoxia, respectively (for more details see Table 2 and case reports in Supplementary Materials). Nine out of ten patients received immunoglobulin replacement therapy in combination with antibiotic and antifungal prophylaxis. Five patients (three SCID, two CID) required HSCT (details on treatment are provided in Supplementary Table 1). Three of the four SCID patients died from infectious and HSCT-related complications before 1-year of age, highlighting the profound immune deficiency with high mortality. In contrast, mortality in the CID cohort was lower, with only one patient (P8) dying between 21-25 years of age due to sepsis with multiorgan failure caused by multiple pathogens (*Stenotrophomonas*, *Pseudomonas*, and Adenovirus). Detailed clinical features and immunological characteristics are provided in Tables 1-3, Supplementary Table 1, and the case reports (Supplementary Materials).

**Figure 1.**
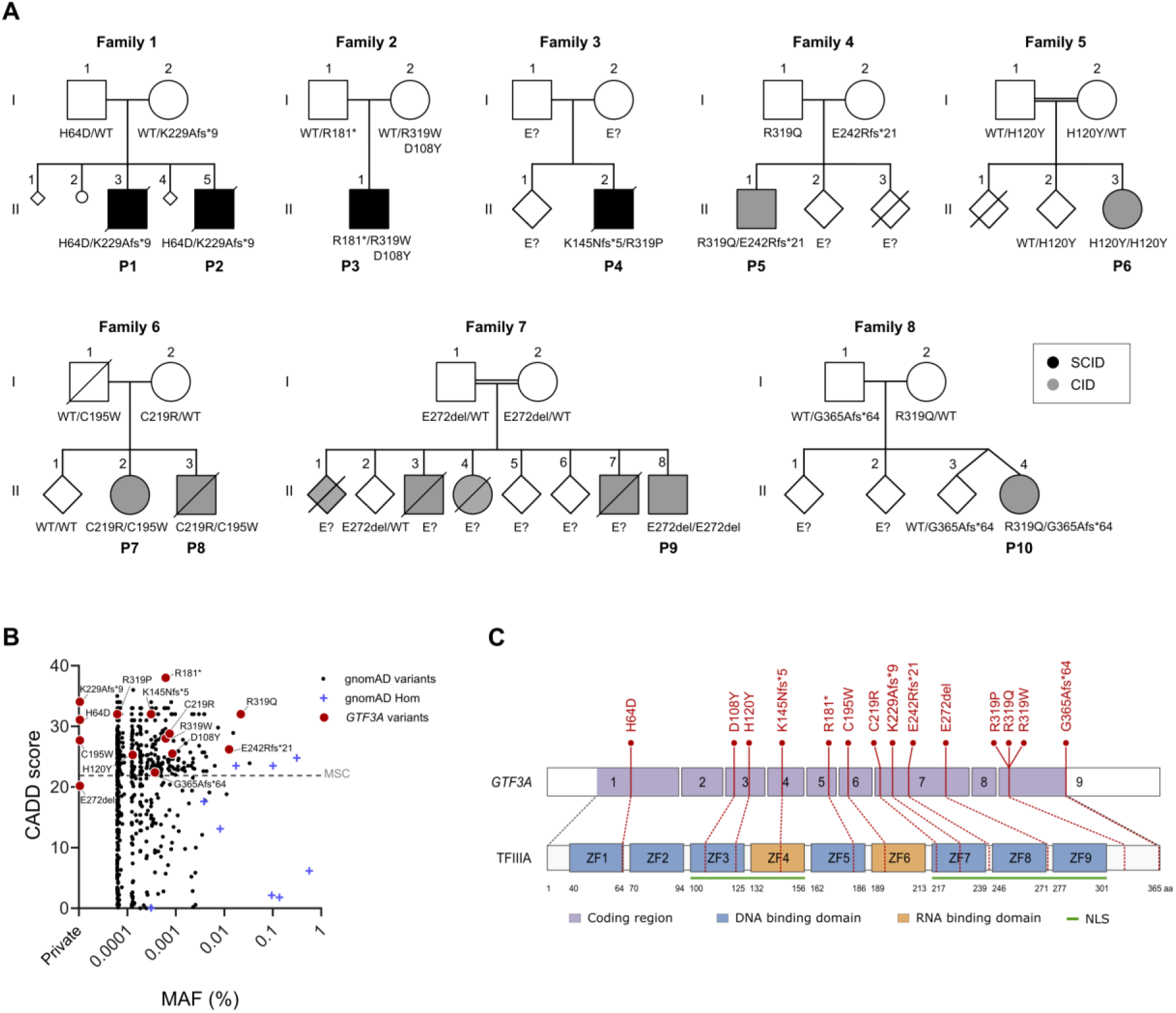
| Identification of GTF3A variants in patients with SCID and CID. (**A**) Pedigrees of eight unrelated families carrying GTF3A variants, showing segregation of alleles with disease. WT denotes wild-type alleles; E? indicates unknown genetic status. (**B**) CADD score plotted against minor allele frequency (MAF) for GTF3A variants reported in gnomAD (v4.1.0), alongside variants identified in our patient cohort. The mutational significance cutoff (MSC; 99th percentile) is indicated by a dotted line at a CADD score of 21.89. (**C**) Schematic representation of the TFIIIA protein, depicting annotated functional domains and the positions of patient-identified variants.

**Table 1.** | Demographic and genetic data of ten TFIIIA-deficient patients from eight unrelated families, all presenting with either SCID or CID.

| Patient no. | P1 | P2 | P3 | P4 | P5 | P6 | P7 | P8 | P9 | P10 |
| --- | --- | --- | --- | --- | --- | --- | --- | --- | --- | --- |
| **Pedigree** | Family 1 | Family 1 | Family 2 | Family 3 | Family 4 | Family 5 | Family 6 | Family 6 | Family 7 | Family 8 |
| **Gender** | Male | Male | Male | Male | Male | Female | Female | Male | Male | Female |
| **Consanguinity** | - | - | - | - | - | + | - | - | + | - |
| **Diagnosis** | **SCID**<br>(T-B<sup>+</sup>NK<sup>+</sup>) | **SCID**<br>(T-B<sup>+</sup>NK<sup>+</sup>) | **SCID**<br>(T-B<sup>+</sup>NK<sup>+</sup>) | **SCID**<br>(T-B<sup>+</sup>NK<sup>+</sup>) | **CID** | **CID** | **CID** | **CID** | **CID** | **CID** |
| **Newborn screening** | - | + | + | + | - | - | - | - | - | - |
| **Current status** | Deceased | Deceased | Alive | Deceased | Alive | Alive | Alive | Deceased | Alive | Alive |
| **Genotype (NM_002097.3)** |  |  |  |  |  |  |  |  |  |  |
| Nucleotide mutations | c.190 C>G/<br>c.682_683dup | c.190 C>G/<br>c.682_683dup | c.541C>T/<br>c.322G>T; c.955C>T | c.956G>C/<br>c.435del | c.956G>A/<br>c.722dup | c.358C>T/<br>c.358C>T | c.585T>G/<br>c.655T>C | c.585T>G/<br>c.655T>C | c.814_816del/<br>c.814_816del | c.956G>A/<br>c.1090del |
| Predicted protein alterations | p.H64D/<br>p.K229Afs*9 | p.H64D/<br>p.K229Afs*9 | p.R181*/<br>p.D108Y; R319W | p.R319P/<br>p.K145Nfs*5 | p.R319Q/<br>p.E242Rfs*21 | p.H120Y/<br>p.H120Y | p.C195W/<br>p.C219R | p.C195W/<br>p.C219R | p.E272del/<br>p.E272del | p.R319Q/<br>p.G365Afs*64 |

**Table 2.**
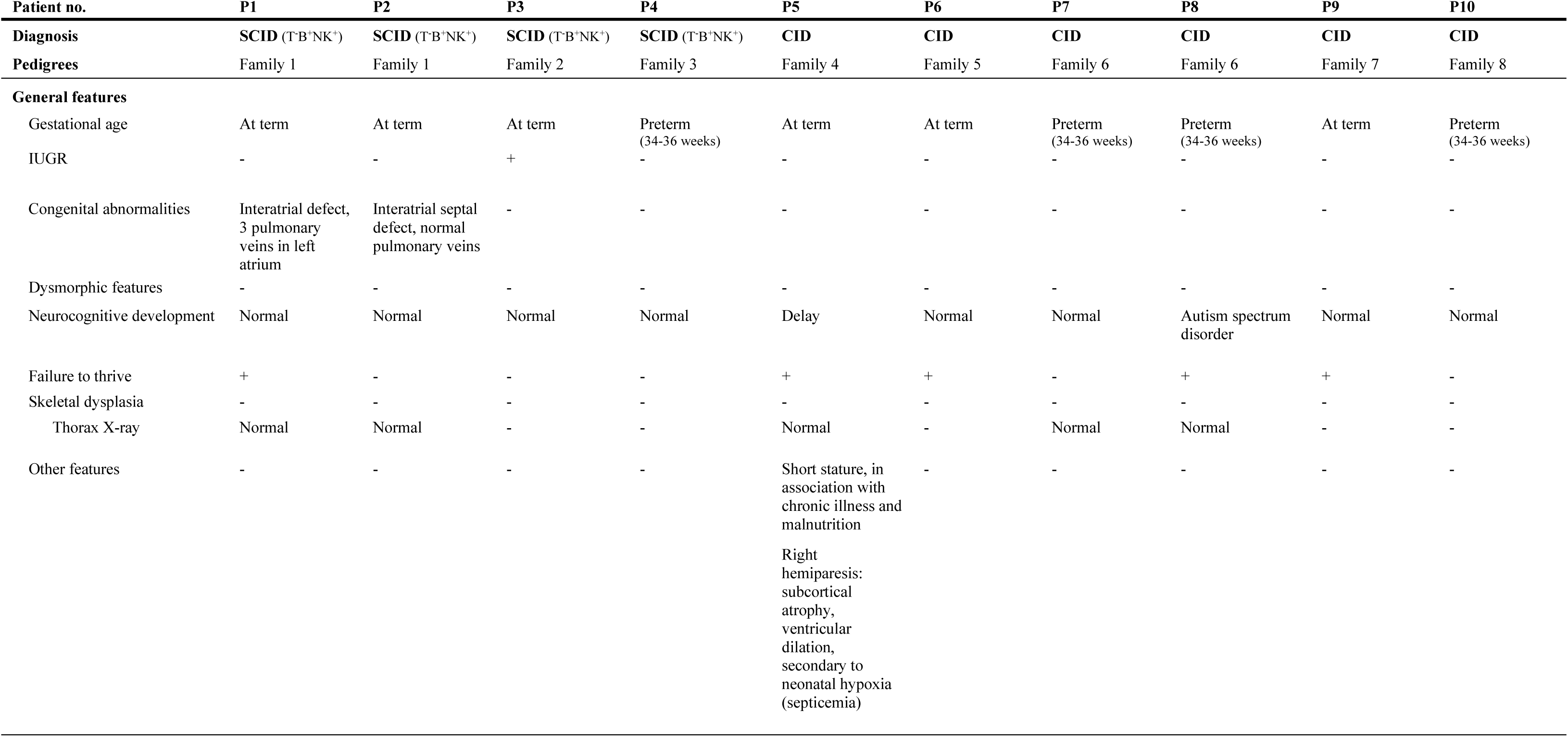
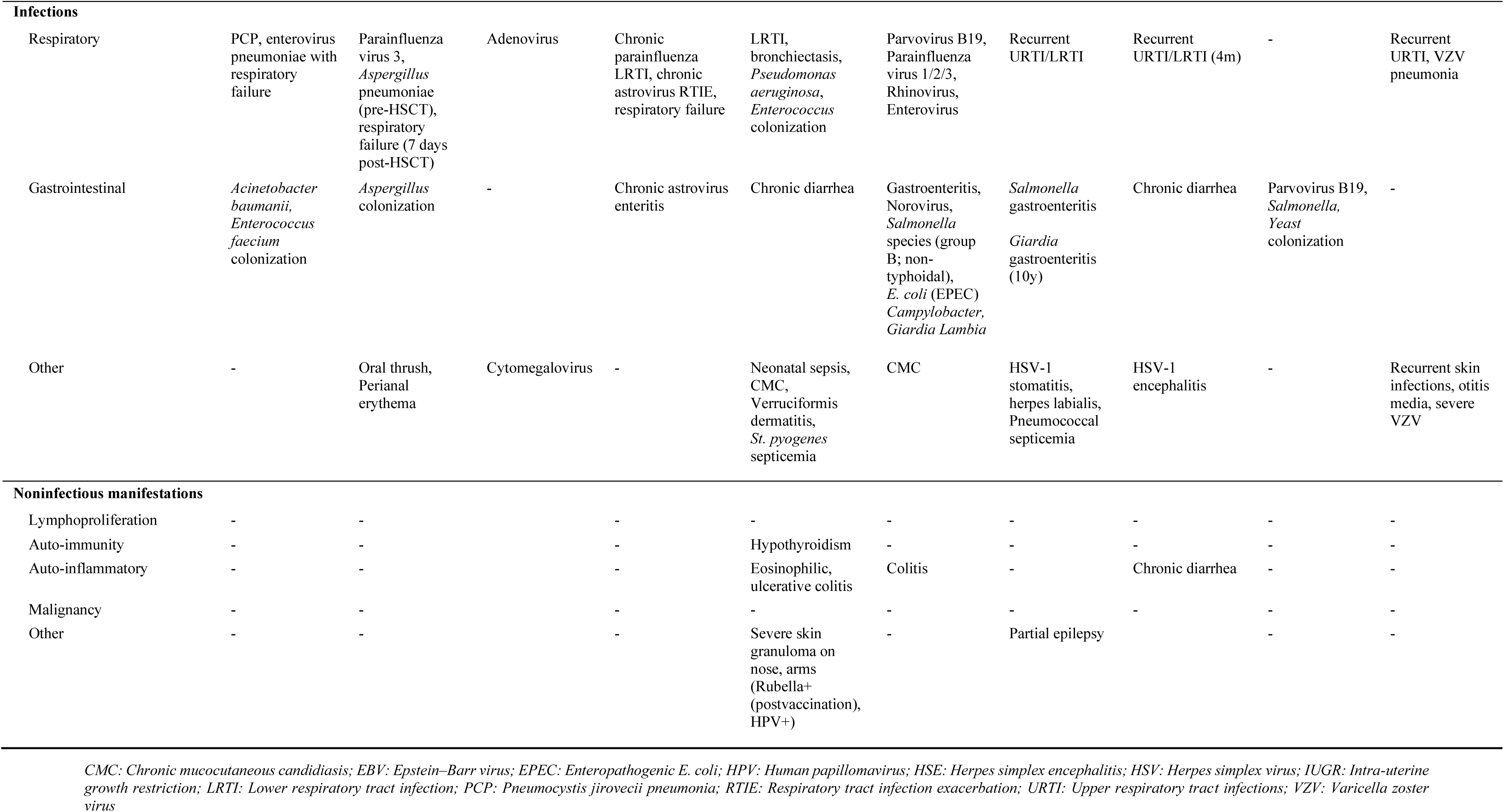
| Clinical characteristics of ten TFIIIA-deficient patients from eight unrelated families. .

**Table 3.**
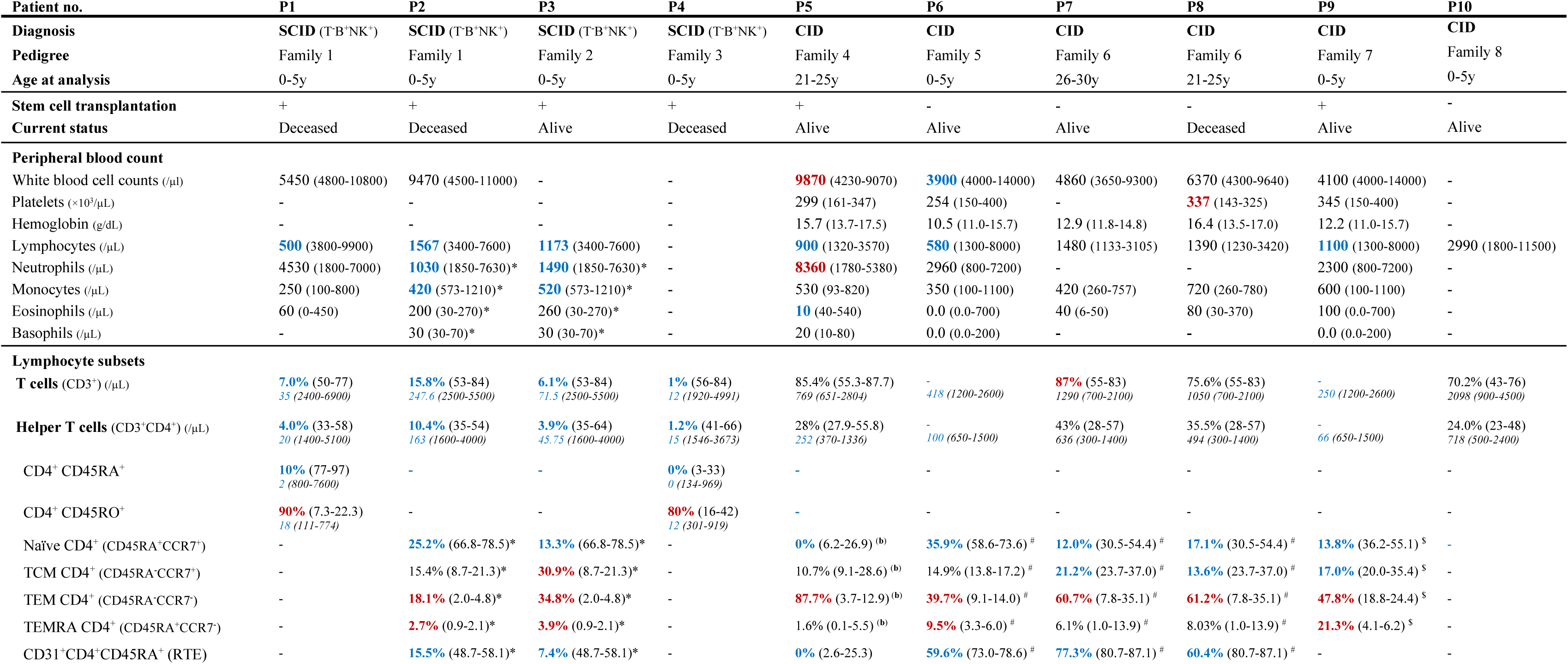
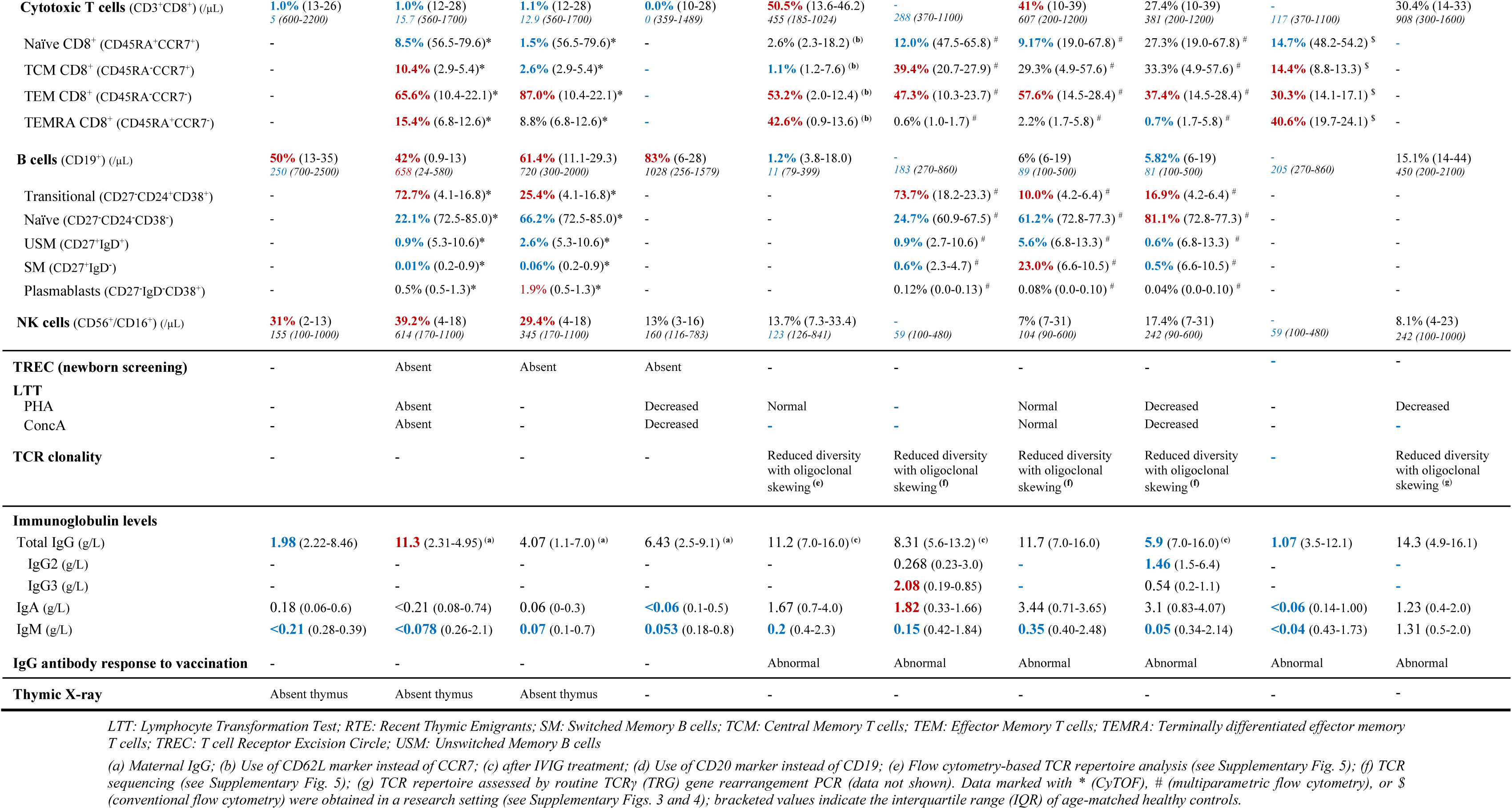
| Immunological profiling of TFIIIA-deficient SCID and CID patients. Immunological data were obtained from both routine clinical testing and research-based analyses using CyTOF immunophenotyping (*), multiparametric flow cytometry (#), or conventional flow cytometry ($). Values highlighted in blue bold indicate reductions, whereas values in red bold indicate elevations relative to age-matched reference ranges (shown in brackets).

Using whole exome sequencing (WES) or whole genome sequencing (WGS), we identified 14 different biallelic variants in the *GTF3A* gene across all patients, segregating with autosomal recessive disease (segregation available in all pedigrees except family 3) (Fig. 1A, Table 1, and Supplementary Table 2). The constraint scores for *GTF3A* indicate a high tolerance for heterozygous loss-of-function (LoF) variants (pLI: 0; LOEUF: 1.0606), but no homozygous LoF variants could be identified in the population database gnomAD v4.1.0 (Supplementary Table 3) (*23*). Indeed, SCoNeS, an integrative metric for negative selection, predicted that *GTF3A* most likely underlies an autosomal recessive inheritance pattern (SCoNeS: 0.918) (*24*). Analysis of gnomAD v4.1.0 indicated that 4 out of the 14 variants were private, while the remaining 10 variants were exclusively observed in a heterozygous state, all with minor allele frequencies (MAF) lower than 0.001 (Fig. 1B and Supplementary Table 2) (*23*).

#### *In silico* evidence supports causality of identified variants

The identified variants included missense substitutions (8/14), frameshift variants (4/14), a nonsense variant (1/14), and an in-frame deletion (1/14), distributed across the zinc finger domains, linker regions, and the C-terminal region of TFIIIA (Fig. 1C and Supplementary Fig. 1A). All 14 variants had high CADD scores exceeding the mutation significance cutoff (MSC; 99% CI), except for the E272del variant, and seven out of eight missense variants affected highly evolutionarily conserved residues (Fig. 1B and Supplementary Fig. 1B) (*25*). Pathogenicity predictions using complementary *in silico* tools, including the meta-predictor REVEL and deep learning-based predictor AlphaMissense, supported the likely deleterious effect for six of the eight missense variants (Supplementary Fig. 1C) (*26*, *27*). Notably, several variants disrupted canonical cysteine or histidine residues essential for C_2_H_2_ zinc finger folding and function, including H64D (in trans with the K229Afs*9; family 1), the homozygous variant H120Y (family 5), and the compound heterozygous variants C195W and C219R (family 6), affecting zinc fingers 1, 3, 6, and 7, respectively (Supplementary Fig. 1B). The nonsense variant R181* (family 2) and frameshift variants K145Nfs*5 (family 3), K229Afs*9 (family 1), and E242Rfs*21 (family 4) are predicted to result in truncated proteins lacking the C-terminal activation domain, previously shown to be required for productive RNA polymerase III-mediated transcription (*28*, *29*). Finally, in four out of eight pedigrees, a missense variant at position R319 (mutated to a proline, glutamine, or tryptophan residue) was identified in the C-terminal region, suggesting a potential mutational hotspot associated with disease. Taken together, the identification of biallelic *GTF3A* variants in multiple independent families, combined with the predicted functional consequences and supporting *in silico* evidence, implicate *GTF3A* as a plausible candidate gene underlying the observed (S)CID phenotype.

#### Reduced TFIIIA protein expression and nascent 5S rRNA in patient-derived fibroblasts

To assess the impact of the identified variants on TFIIIA protein expression, we performed western blot analysis in available patient-derived fibroblasts. Antibody specificity was confirmed using siRNA-mediated *GTF3A* knockdown in human fibroblasts (Supplementary Fig. 2A). Consistent with predicted LOF effects, fibroblasts from patients carrying truncating variants, *in trans* with missense variants (P2, H64D/K229Afs*9; P5, R319Q/E242Rfs*21; and P3, R181*/D108Y;R319W), showed markedly reduced TFIIIA protein levels compared to healthy controls (Fig. 2A). Reduced protein expression was also observed in fibroblasts from two siblings carrying compound heterozygous missense variants (P7 and P8, C195W/C219R).

**Figure 2.**
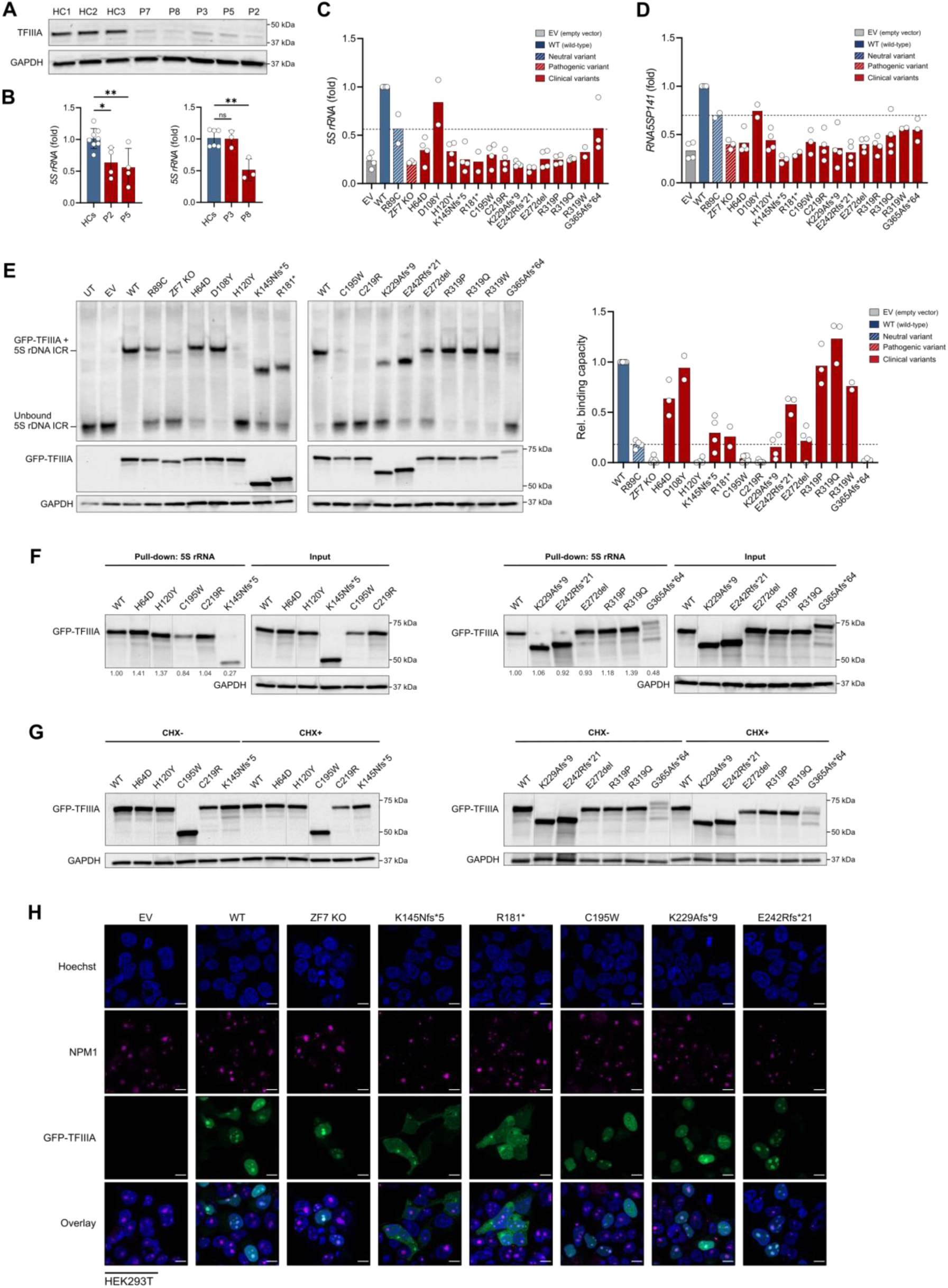
| Disease-associated *GTF3A* variants disrupt multiple TFIIIA molecular functions. (**A**) Endogenous TFIIIA protein expression in fibroblasts from HCs, P2, P3, P5, P7, and P8, assessed by immunoblotting. Western blot is representative of two independent experiments. (**B**) RT-qPCR analysis of 5S rRNA expression in primary fibroblasts from patients (P2, P3, P5, and P8) compared to healthy controls (HCs, n=2). Pooled data from at least three independent repeats is shown. (**C**) RT-qPCR analysis of 5S rRNA expression in TFIIIA C195W/C219R knock-in (KI) HEK293T cells that were transfected for 16 hours with empty vector (EV), GFP-tagged wild type (WT), or mutant TFIIIA. Data are expressed as fold change relative to WT-rescued cells. R89C serves as a functional reference: a gnomAD variant with partial TFIIIA activity but no known clinical significance; the dashed line indicates its activity threshold. Pooled data from at least two independent repeats per variant is shown. (**D**) RT-qPCR analysis of *RNA5SP141* expression in TFIIIA C195W/C219R KI HEK293T cells that were transfected for 16 hours with EV, GFP-tagged WT, or mutant TFIIIA. Data are expressed as fold change relative to WT-rescued cells. Pooled data from at least two independent repeats per variant is shown. (**E**) Electrophoretic mobility shift assay (EMSA) showing TFIIIA binding to the *5S rDNA* internal control region (ICR) in WT HEK293T cells transfected with EV, GFP-tagged WT, or mutant TFIIIA constructs. Representative EMSA images are shown with densitometric quantification from at least two independent experiments per variant. The R89C variant indicates a reference level for reduced DNA-binding activity. (**F**) TFIIIA binding to 5S rRNA assessed by RNA-pull down in HEK293T cells transfected with either EV, GFP-tagged WT, or mutant TFIIIA constructs. Values below the blot represent pull-down/input ratios normalized to WT. (**G**) Cycloheximide (CHX)-chase assay (24h) performed on WT HEK293T cells transfected with the indicated GFP-tagged WT or mutant TFIIIA, demonstrating TFIIIA protein stability. (**H**) Confocal imaging of WT HEK293T cells transfected with either EV, GFP-tagged WT, or mutant TFIIIA constructs. Representative immunofluorescence images are shown, with Hoechst and nucleophosmin (NPM1) stainings marking the nucleus and nucleolus, respectively. Scale bars are 10 µm. Data in (E-H) are representative for at least two independent repeats. Statistical test: One-way ANOVA with Dunnett’s multiple comparisons (B). ns: not significant, *P<0.05, **P<0.01.

Given the central role of TFIIIA in mediating transcription of 5S rRNA, we next evaluated whether patient-derived *GTF3A* variants impair this core molecular function in primary cells. Steady-state 5S rRNA levels were significantly reduced in fibroblasts derived from patients P2, P5, and P8, while P3 demonstrated levels comparable to controls (Fig. 2B). To more directly evaluate TFIIIA-mediated transcriptional output, we labeled newly synthesized RNA with 5-ethynyl uridine (EU) (*30*). In line with steady-state results, P2, P5, and P8 demonstrated reduced newly synthesized 5S rRNA levels, albeit mildly reduced in P5 (Supplementary Fig. 2B). EU labeling revealed a marked reduction in nascent 5S rRNA in P3, indicative of a defect in 5S rRNA transcriptional kinetics masked at the steady-state level by the high abundance and stability of 5S rRNA (Supplementary Fig. 2B).

### *GTF3A* variants disrupt 5S rRNA and *RNA5SP141* pseudogene transcription

To systematically assess the impact of all identified *GTF3A* variants, we next established an orthogonal overexpression-based assay using a knock-in (KI) HEK293T cell line engineered to carry the previously validated LOF variants C195W and C219R (family 6) (*21*). KI HEK293T cells maintained steady-state 5S rRNA levels comparable to WT cells, yet exhibited significantly reduced nascent 5S rRNA upon EU labeling (Supplementary Fig. 2C). In this system, transient overexpression of wild-type (WT) TFIIIA robustly increased 5S rRNA levels, whereas a ZF7 knockout (KO) construct, previously shown to lack DNA-binding capacity (*21*), failed to increase expression levels, comparable to the empty vector (EV) control (Fig. 2C, Supplementary Fig. 2D). With the exception of the D108Y variant (present in *cis* with R319W and in *trans* with R181* in P3), all patient-derived *GTF3A* variants showed markedly reduced TFIIIA-mediated 5S rRNA transcription, remaining well below WT levels despite efficient expression of the corresponding TFIIIA proteins (Fig. 2C and Supplementary Fig. 2D).

To contextualize these defects, we included the homozygous R89C variant, present in gnomAD v4.1.0 and previously shown to partially impair DNA binding (*21*), as a functional reference for reduced TFIIIA activity without known clinical ramifications. As expected, overexpression of R89C mediated only a partial increase in 5S rRNA transcription and did not reach WT activity (Fig. 2C and Supplementary Fig. 2D). With the exception of D108Y and the G365Afs*64 variants, none of the remaining patient-derived *GTF3A* variants restored 5S rRNA transcription above the R89C threshold, indicating a convergent reduction in TFIIIA-mediated transcriptional activity across the cohort.

In addition to its role in canonical 5S rRNA transcription, TFIIIA regulates the expression of a large family of *5S rRNA* pseudogenes (*31*). Among these, the pseudogene *RNA5SP141* has been shown to play a critical role in innate antiviral immunity by acting as an endogenous ligand for the cytosolic sensor RIG-I, thereby promoting the induction of antiviral cytokine responses to a broad range of DNA and RNA viruses, including herpesviruses(*21*, *22*). To determine whether the observed TFIIIA transcriptional defect extends beyond canonical 5S rRNA, we next quantified *RNA5SP141* expression using the same overexpression-based KI HEK293T system. Consistent with the impaired TFIIIA function in these cells, KI HEK293T cells exhibit reduced *RNA5SP141* expression compared to WT HEK293T cells, underscoring the dependence of *RNA5SP141* transcription on intact TFIIIA activity (Supplementary Fig. 2E). Thirteen out of fourteen patient-derived *GTF3A* variants failed to restore *RNA5SP141* expression to WT levels (Fig. 2D), closely mirroring their effects on canonical 5S rRNA transcription. In contrast, the D108Y variant (present in *cis* with R319W in P3) restored *RNA5SP141* expression to levels comparable to R89C, supporting the notion that this allele retains a degree of TFIIIA transcriptional activity and might thus be functionally neutral.

#### Disruption of zinc fingers 3, 6, and 7 impair ICR binding by TFIIIA

The observed defects in TFIIIA-mediated 5S rRNA and *RNA5SP141* transcription prompted us to investigate the underlying molecular mechanisms. As TFIIIA initiates transcription through direct interaction with the internal control region (ICR) of *5S rDNA* genes, we assessed whether patient-derived *GTF3A* variants retain ICR-binding capacity using an electrophoretic mobility shift assay (EMSA) (*20*, *32*). GFP-tagged WT and mutant TFIIIA constructs were overexpressed in WT HEK293T cells, and their ability to bind an ICR-containing DNA probe was evaluated.

Consistent with prior studies in *Xenopus* and yeast identifying ZF1-3, 5, and 7-9 as core ICR-binding domains of TFIIIA (*20*, *32–34*) (Fig. 1C), variants affecting these regions showed reduced DNA-binding activity compared to WT TFIIIA, with the exception of the D108Y variant (in line with its retained transcriptional activity) (Fig. 2E). In particular, the H120Y variant (ZF3) and the C219R variant (ZF7) displayed near-complete loss of ICR binding, comparable to a ZF7 knockout construct known to lack DNA-binding capacity (Fig. 2E) (*21*). Using the R89C variant as a functional reference for partial yet clinically tolerated impairment of ICR binding, we observed that four variants, K145Nfs*5 (ZF4-9), R181* (ZF5-9), K229Afs*9 (ZF7-9), and E272del (linker region), clustered near this reference level, consistent with a modest impairment of DNA-binding capacity (Fig. 2E). Conversely, two additional variants, H64D (ZF1) and E242Rfs*21 (ZF8-9), retained ICR-binding well above the R89C threshold despite reduced binding relative to WT TFIIIA (Fig. 2E). This partial preservation of DNA binding was observed even though these variants affect ZFs defined as critical for ICR recognition, suggesting that their pathogenic effects on TFIIIA-mediated transcription are unlikely to be driven solely from loss of ICR binding. The C195W variant (ZF6) and the G365Afs*64 variant (C-terminal) displayed markedly reduced DNA binding, despite lying outside previously defined ICR-binding domains, suggesting that these specific alterations may indirectly destabilize TFIIIA-DNA interactions (Fig. 2E). All variants affecting residue R319 (R319P, R319Q, and R319W) retained ICR-binding capacity comparable to WT TFIIIA, consistent with their location in the C-terminal region (Fig. 2E).

#### RNA-binding and TFIIIA protein stability defects associated with selected *GTF3A* variants

Beyond its role in *5S rDNA* transcription, TFIIIA functions as a chaperone by directly binding nascent 5S rRNA, thereby promoting its stability and facilitating proper subcellular trafficking toward nucleolar ribosome assembly sites (*16*, *19*). To assess whether this RNA-binding function is disrupted by patient-derived *GTF3A* variants, we performed an RNA pull-down assay using biotinylated 5S rRNA as bait. Both variants located in ZF4 and ZF6, K145Nfs*5 and C195W, showed markedly reduced RNA-binding capacity compared with WT TFIIIA (Fig. 2F), consistent with prior studies identifying these ZFs as critical mediators of 5S rRNA binding and chaperoning (*16*, *17*, *19*). The C-terminal G365Afs*64 variant also displayed reduced RNA binding, while the remaining patient-derived variants retained RNA-binding capacity comparable to WT TFIIIA (Fig. 2F).

Of note, the C-terminal G365Afs*64 variant demonstrated dual impairment in both DNA and RNA binding, even though the mutation resides outside the established binding domains (Fig. 2E, F). Given that this frameshift variant introduces an aberrant C-terminal extension of 64 amino acids, we hypothesized that the observed binding defects might arise from protein misfolding and consequent instability, rather than direct disruption of functional binding domains. To test this, we evaluate TFIIIA turnover using a cycloheximide (CHX) chase assay. While all patient-derived variants, including truncated proteins, maintained stability comparable to WT protein, the G365Afs*64 mutant showed pronounced instability, indicating that the functional impairments are likely driven by protein instability or conformational disruption (Fig. 2G and Supplementary Fig. 2F).

#### Impaired nuclear import of TFIIIA upon NLS disruption by truncating variants

The molecular functions of TFIIIA critically depend on its subcellular localization. TFIIIA is synthesized in the cytoplasm and subsequently imported into the nucleus via defined nuclear localization signal (NLS) regions. Two NLS regions have been described based on studies in *Xenopus*: an NLS spanning ZFs 3-4 and a second NLS encompassing ZFs 7-9 (Fig. 1C) (*35*). Following nuclear entry, TFIIIA mediates 5S rRNA transcription in the nucleoplasm and subsequently binds 5S rRNA to facilitate its trafficking to the nucleolus for ribosome biogenesis. Although less characterized, TFIIIA-5S rRNA complexes have also been reported to shuttle back to the cytoplasm, suggesting additional regulatory roles (*35*, *36*).

As several patient-derived *GTF3A* variants are predicted to disrupt one or both NLS regions, we hypothesized that altered subcellular localization could contribute to TFIIIA dysfunction. To test this, we performed confocal microscopy in HEK293T cells transiently expressing N-terminally GFP-tagged WT or mutant TFIIIA constructs, focusing on variants predicted to affect NLS integrity. WT TFIIIA localized predominantly to the nucleus, with pronounced nucleolar enrichment as confirmed by Nucleophosmin 1 (NPM1) co-staining (Fig. 2H). Variants predicted to severely disrupt NLS integrity exhibited clear defects in nuclear import. The K145Nfs*5 variant, which truncates TFIIIA within ZF4 and disrupts both predicted NLS regions, showed marked cytoplasmic accumulation with minimal nuclear localization (Fig. 2H). Similarly, the R181* variant, predicted to disrupt the full C-terminal NLS (ZF7-9), accumulated predominantly in the cytoplasm. In contrast, variants predicted to only partially affect the second NLS retained normal localization. Two frameshift variants, K229Afs*9 and E242Rfs*21, which truncate TFIIIA downstream of ZF7 and partially disrupt the second NLS region, displayed nuclear localization patterns comparable to WT TFIIIA. Consistent with these observations, the ZF7 KO construct also retained normal nuclear localization (Fig. 2H).

Because the K145Nfs*5 and R181* variants impair or are predicted to impair TFIIIA’s RNA binding capacity (Fig. 2F), we sought to determine whether the observed mislocalization reflected disruption of NLS sequences rather than secondary effects of defective RNA binding. To address this, we included the non-truncating C195W variant (ZF6), which markedly reduces RNA-binding capacity but does not disrupt the predicted NLS regions. Notably, C195W retained efficient nuclear localization comparable to WT TFIIIA, indicating that impaired RNA binding alone does not perturb TFIIIA nuclear import (Fig. 2H). Together, these results establish defective nuclear import due to NLS disruption as an additional pathogenic mechanism in TFIIIA deficiency, thereby limiting TFIIIA access to nuclear DNA targets and likely contributing to the observed transcriptional alterations.

#### TFIIIA deficiency causes profound T cell lymphopenia across SCID and CID presentations

Next, we performed detailed immunophenotyping to define shared and distinct immune features across the SCID-CID spectrum of TFIIIA deficiency. Across both SCID and CID presentations, TFIIIA deficiency was characterized by a consistent and predominant T cell defect. All SCID patients exhibited markedly reduced percentage of CD3⁺ T cells affecting both CD4⁺ and CD8⁺ subsets, while CID patients showed consistently reduced CD4⁺ T cell numbers with variable loss of CD8⁺ T cells (Table 3). Comprehensive immune profiling by mass cytometry (CyTOF) in two SCID patients (P2 and P3) and multiparametric flow cytometry in five CID patients (P5-P9) corroborated these findings and further revealed a profound decrease in naïve CD4⁺ and CD8⁺ T cell subsets in both phenotypes, accompanied by relative enrichment of central and/or effector memory T cell subsets (Table 3, Fig. 3A-D, Supplementary Fig. 3A, B). CD31⁺ recent thymic emigrants (RTEs) were consistently low to absent across all patients, concordant with reduced to absent TREC levels (Table 3, Fig. 3E) (*37*). Tregs were increased in relative frequency in SCID patients, whereas they were normal to moderately decreased in CID patients (Fig. 3F). Of note, siblings P7 and P8, and P10 demonstrated a milder phenotype, with low to normal total CD3⁺, CD4⁺, and CD8⁺ T cell counts, but a consistently contracted naïve compartment compared to age-matched reference ranges (Table 3).

**Figure 3.**
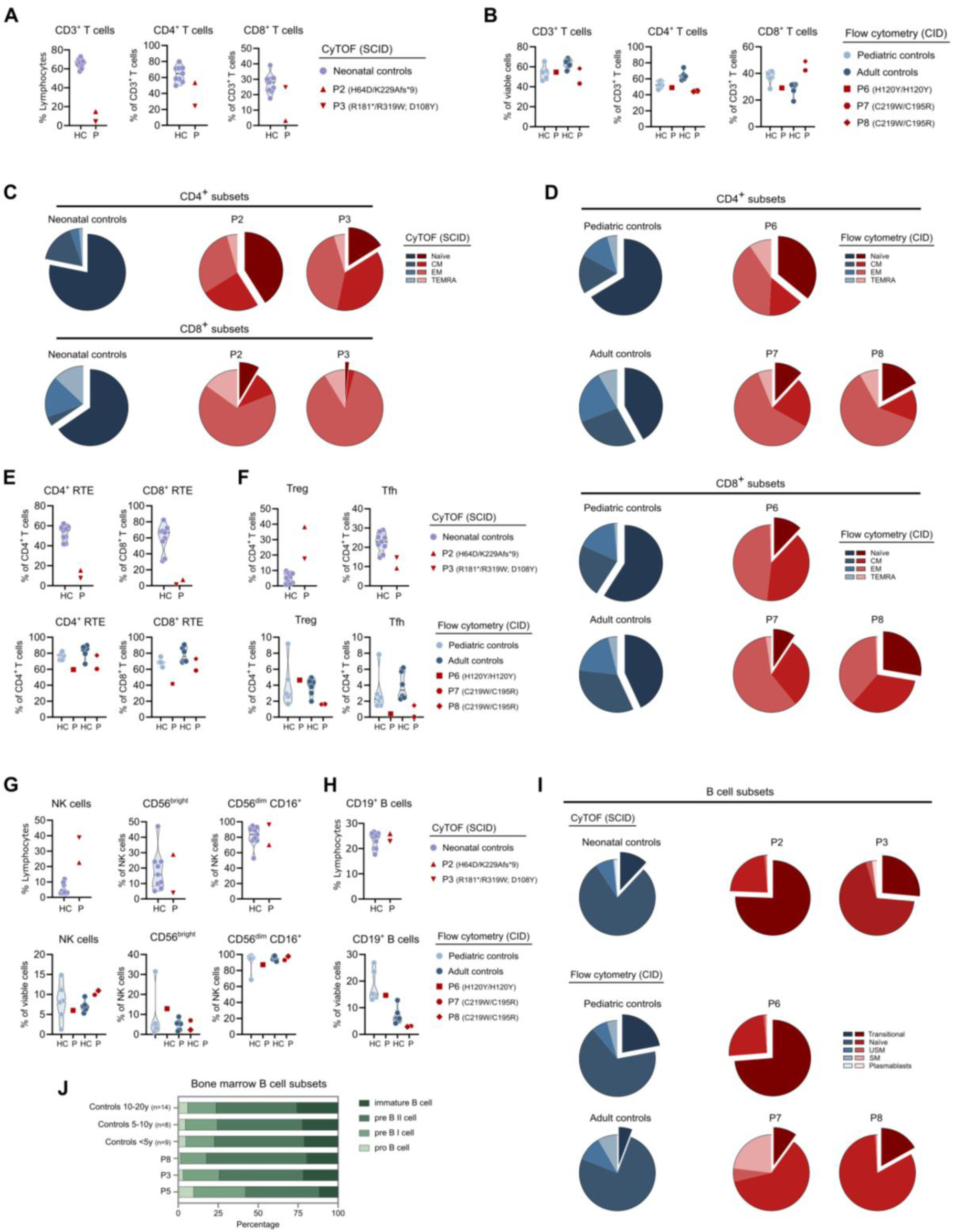
| Immunophenotyping identifies a T⁻B⁺NK⁺ SCID or CID phenotype with predominant T cell lymphopenia. (**A-F**) Frequencies of T cell subsets in whole blood or peripheral blood mononuclear cells (PBMCs) from two TFIIIA-deficient SCID patients (P2, P3) and three CID patients (P5, P6, P7), respectively. Age-matched neonatal (n=9), pediatric (n = 6), and adult controls (n = 6) were included. SCID patients were analyzed by Cytometry by Time of Flight (CyTOF); CID patients were analyzed by 27-parameter multiparametric flow cytometry with manual gating. T cell subsets were defined as naïve (CCR7^+^CD45RA^+^), central memory (CM; CCR7^+^CD45RA^−^), effector memory (EM; CCR7^−^CD45RA^−^), terminally differentiated effector memory (TEMRA; CCR7^−^CD45RA^+^), recent thymic emigrants (RTE; CCR7^+^CD45RA^+^CD31^+^), regulatory T cells (Treg; CD25^+^CD127^-^), and T follicular helper (Tfh; CXCR5^+^CD95^+^). (**G**) NK cell frequencies in SCID and CID patients and age-matched controls, determined by either CyTOF or flow cytometry. (**F-G**) Total B cell frequency and B cell subset distribution in SCID, patients, CID patients, and age-matched controls, analyzed by either CyTOF or flow cytometry. B cell subsets were defined as transitional (CD27^-^CD24^+^CD38^+^), naïve (CD27^-^CD24^-^CD38^-^), unswitched memory (USM; CD27^+^IgD^+^), switched memory (SM; CD27^+^IgD^-^), and plasmablasts (CD27^-^IgD^-^CD38^+^). (**J**) Frequencies of B cell precursor subsets in bone marrow from a SCID patient (P3) and two CID patients (P5, P8), compared with age-matched controls (<5 years, n=9; 5-10 years, n=8; 10-20 years, n=14). B cell precursor subsets in P3 and P8 were defined as pro B cells (CD34⁺CD19⁻TdT^+^), pre-B I cells (CD19⁺cyIgM⁻), pre-B II cells (CD19⁺cyIgM⁺), and immature B cells (CD19⁺sIgM⁺IgD^-^). B cell precursor subsets in P5 were defined as pro B cells (CD34^+^CD19^-^CD22^+^), pre-B I cells (CD34^+^CD19^+^), pre-B II cells (CD19⁺CD34^-^CD20^-or^ ^dim^CD10^+^), and immature B cells (CD19⁺CD34^-^CD20^+^CD10^+^). Data (A-O) are representative of a single experiment.

Analysis of unconventional T cell subsets revealed largely preserved mucosal-associated invariant T (MAIT) cell frequencies across both groups, whereas γδ T cells were mainly increased in SCID patients (P2 and P3). Invariant natural killer T (iNKT) cell frequencies were normal in the SCID groups, while in the setting of CID, they were reduced in childhood but normal in adults (Supplementary Fig. 3C). TCR repertoire analyses performed in four CID patients using flow cytometry (P5) or high-throughput TCR sequencing (P6, P7, P8) revealed reduced TCR diversity with skewing towards oligoclonality across all patients (Table 3, Supplementary Fig. 4A-D). Despite this overall restriction, the frequency of TCR Vα7.2⁺ CD3⁺ T cells was preserved, indicating that TCRα rearrangements associated with MAIT cell development are maintained (Supplementary Fig. 4E).

In contrast to the observed T cell defects, NK cell compartments were largely intact across both SCID and CID groups, with normal absolute counts and intact subset distribution, including CD56^bright^ and CD56^dim^CD16⁺ NK cell populations (Table 3; Fig. 3G). The myeloid compartment was similarly unaffected, with only modest reductions in neutrophils and monocytes in P2 and P3, while eosinophil and basophil counts remained within normal ranges. (Table 3, Supplementary Fig. 3D).

#### B cell compartment abnormalities and humoral immune deficiency are more prominent in CID

The phenotype of the B cell compartment was more heterogeneous, exhibiting distinct features across the SCID-CID spectrum. In SCID patients, B cell percentages were preserved or elevated relative to age-matched reference ranges, consistent with a T^-^B^+^NK^+^ immunophenotype (Table 3, Fig. 3H). CyTOF analysis in P2 and P3 revealed increased transitional B cells, with reductions in naïve and memory B cell subsets including both unswitched and switched memory B cells, while plasmablast frequencies remained comparable to neonatal controls (Table 3, Fig. 3I, Supplementary Fig. 3E). In contrast, absolute B cell counts were moderately reduced in five out of six CID patients, with variably reduced relative B cell frequencies (Table 3, Fig. 3H). More detailed analysis in three CID patients (P6, P7, P8) demonstrated skewing towards increased transitional B cells, with variably affected naïve B cells and reductions in unswitched and switched memory B cell subsets in P6 and P8, and relative expansion in P7 (Table 3, Fig. 3I, Supplementary Fig. 3E). Plasmablast frequencies were comparable to controls. In parallel, circulating Tfh cells, critical regulators of germinal center formation and B cell maturation, were consistently reduced across both SCID and CID patients (Fig. 3F).

To investigate whether the observed B cell abnormalities reflected impaired early B cell development, we analyzed bone marrow B cell precursors in one SCID patient (P3) and two CID patients (P5 and P8) (*38*). Flow cytometric analysis of pro-B, pre-B I, pre-B II, and immature B cell populations revealed no major differences compared with age-matched controls (Fig. 3J), indicating preserved early B cell development in TFIIIA deficiency.

Humoral immune deficiency was a consistent feature across the cohort. Serum immunoglobulin evaluation revealed uniformly reduced IgM levels in both SCID and CID patients, with the exception of P10, whereas additional reductions in IgG and/or IgA were present in a subset of patients (Table 3). Surface IgM expression on B cells, assessed in two CID patients (P7 and P8), was comparable to healthy controls (Supplementary Fig. 3F). In addition, all evaluated CID patients demonstrated impaired vaccine-specific antibody responses, particularly against polysaccharide antigens including pneumococcal and *Haemophilus influenzae* type B vaccines. No circulating autoantibodies were detected in any patient, arguing against overt humoral immune dysregulation (data not shown). Collectively, these findings define TFIIIA deficiency-associated SCID as a T^-^B^+^NK^+^ immunophenotype, while both the T and B cell compartment and antigen-specific humoral immunity are impaired in patients with TFIIIA deficient CID.

#### TFIIIA is required for early human thymocyte development

Across both SCID and CID patients, immunophenotyping consistently revealed marked reductions in naïve T cells and CD31⁺ RTEs, suggesting impaired thymic T cell output rather than peripheral T cell loss. To directly assess whether TFIIIA deficiency affects human T cell development, we next examined thymocyte differentiation using the artificial thymic organoid (ATO) system. This *in vitro* platform supports stepwise differentiation of human T cells from CD34⁺ hematopoietic progenitors and enables precise determination of the developmental stage at which T cell generation is arrested (Fig. 4A, Supplementary Figure 5A) (*39*). ATO cultures were established from either PBMC or bone marrow-derived CD34⁺ cells from six patients across five families (P2, P3, P5, P6, P7, and P8).

**Figure 4.**
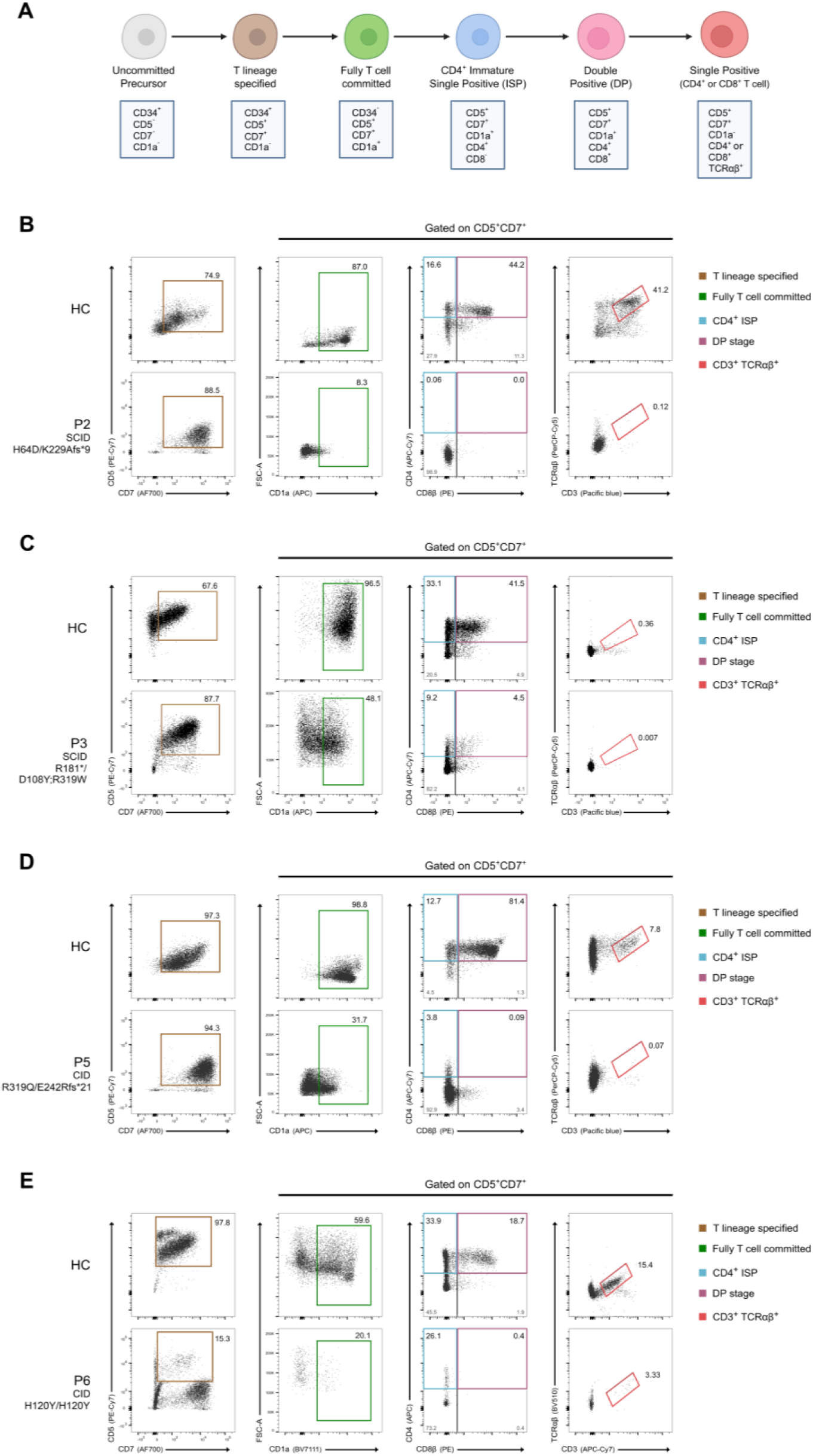
| TFIIIA deficiency causes an early block in human T cell development. (**A**) Schematic overview of human T cell developmental stages supported by the artificial thymic organoid (ATO) system. (**B-E**) *In vitro* T cell differentiation of CD34⁺ hematopoietic progenitors from TFIIIA-deficient patients using the ATO platform: (**B**) patient P2 (SCID; analyzed at 6 weeks), (**C**) patient P3 (SCID; analyzed at 5 weeks), (**D**) patient P5 (CID; analyzed at 5 weeks), and (**E**) patient P6 (CID; analyzed at 5 weeks). Flow cytometry plots show expression of CD7, CD5, CD1a, CD4, CD8β, TCRαβ, and CD3 upon gating on LIVE/DEAD^-^CD45^+^CD56^-^ cells, demonstrating an early developmental arrest in TFIIIA-deficient patients. Data (B-D) are representative of a single experiment per patient.

Across all patient-derived ATO cultures, absolute numbers of CD45⁺CD56^-^ early lymphoid progenitors were markedly reduced compared with healthy controls, indicating impaired survival and/or proliferation at early stages of lymphoid development. Despite this quantitative deficit, residual progenitors in most patients retained the capacity to initiate T-lineage specification and progressed to the CD5⁺CD7⁺ stage (Fig. 4B-E and Supplementary Fig. 5B). Patient P6 was distinguished by a pronounced reduction at the CD5⁺CD7⁺ stage compared with both a healthy control and other TFIIIA-deficient patients, suggesting a more pronounced defect in early T-lineage differentiation (Fig. 4D).

Further analysis of early thymocyte differentiation revealed markedly impaired progression of patient-derived precursors to the T lineage committed CD1a⁺ stage (Fig. 4B-E and Supplementary Fig. 5B). The SCID patient P2 displayed an almost complete block prior to the immature single positive (ISP; CD4⁺CD8β^-^) stage (Fig. 4B). In contrast, SCID patient P3 and CID patients P5 and P6 generated a minimal ISP thymocyte population but failed to produce double positive (DP; CD4⁺CD8β⁺) cells (Fig. 4C-E). Siblings from family 6 (P7 and P8) progressed to the ISP stage and generated a small DP subset, albeit with reduced frequencies compared with controls (Supplementary Fig. 5B). Across all ATO cultures, mature TCRαβ⁺ CD3⁺ T cells were minimally generated, with the exception of P7, who retained a small residual population (Supplementary Fig. 5B). These experiments unambiguously reveal a thymocyte-intrinsic defect at a very early stage in T cell development, prior to V(D)J recombination-associated developmental blocks (*40*).

#### Impaired early T cell development in *gtf3aa*-deficient zebrafish

The early block in human thymocyte differentiation identified in the ATO system indicated that TFIIIA is essential for early T cell development. To determine whether this requirement is conserved *in vivo*, we used the zebrafish model, which enables direct visualization of thymic T cell development using a fluorescent reporter. We employed the *Tg(lck:EGFP)* transgenic zebrafish line, in which EGFP expression driven by the T-lineage-specific protein tyrosine kinase (*lck*) promoter labels both immature and mature T cells from early larval stages through adulthood (*41*).

Owing to a teleost-specific genome duplication, zebrafish harbor two ohnologs of the human *GTF3A* gene, namely *gtf3aa* and *gtf3ab* (*42*). Comparative protein sequence analysis revealed evolutionary conservation, with human TFIIIA sharing 45% amino acid identity with zebrafish *gtf3aa* and 38% with *gtf3ab* (Supplementary Fig. 6A). To assess the *in vivo* requirement for TFIIIA during early thymic T cell development, we performed CRISPR/Cas9-based gene disruption of *gtf3aa*, *gtf3ab,* and *gtf3aa;gtf3ab* and analyzed the F0 (crispant) zebrafish in the *Tg(lck:EGFP)* background.

As *lck* expression is initiated after T cell progenitors colonize the thymus, EGFP-positive cells become detectable from approximately 3 days post-fertilization (dpf), marking the onset of thymic T cell development in zebrafish (*41*, *43*). To evaluate early T cell development, we assessed zebrafish larvae at 5 and 8 dpf. *In vivo* imaging of the thymic region at 5 dpf revealed a significant reduction in the thymic EGFP signal in both *gtf3aa* and *gtf3aa;gtf3ab* crispants compared to control larvae (Fig. 5A, B). This defect persisted at 8 dpf (Supplementary 6B, C), indicating a sustained impairment in early T cell development. In contrast, *gtf3ab* crispants exhibited thymic EGFP expression comparable to controls at both timepoints. These findings are consistent with previous studies identifying *gtf3aa* as the predominantly expressed ohnolog in somatic tissues, whereas *gtf3ab* expression is largely restricted to oocytes (*42*).

**Figure 5.**
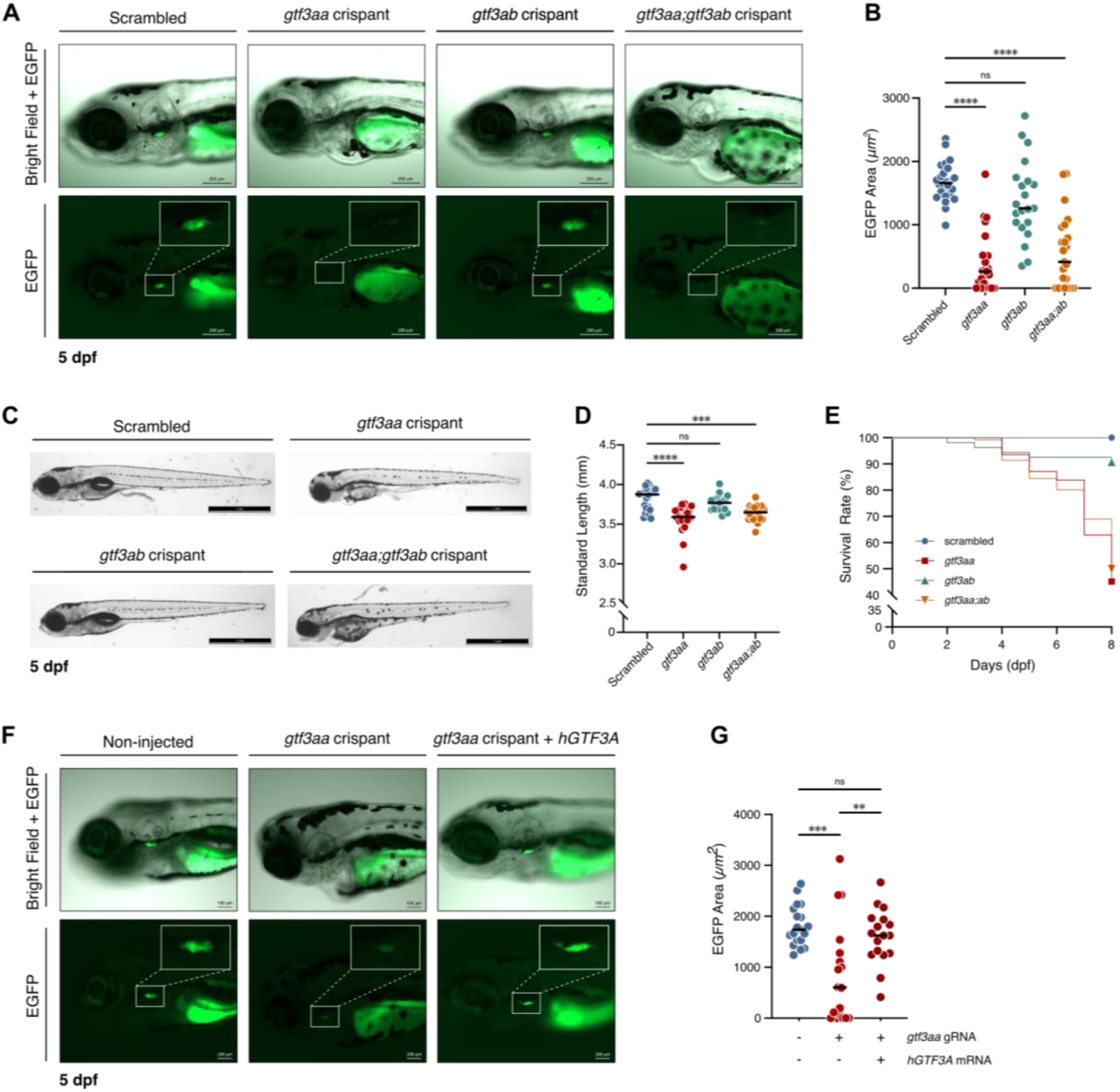
| Loss of *gtf3aa* impairs early T cell development and reduces survival in zebrafish larvae. (**A**) Representative merged bright-field and EGFP fluorescence images, and corresponding EGFP-only images, of the thymic region in Tg(*lck*:EGFP) zebrafish larvae at 5 days post-fertilization (dpf), showing reduced thymic EGFP signal in *gtf3aa* and *gtf3aa;gtf3ab* crispants. White boxes indicate the thymic region. (**B**) Quantification of the EGFP-positive area within the thymic region, defined as the area located below the otic vesicle and above the heart. (**C**) Representative bright-field images of *scrambled* control, *gtf3aa*, *gtf3ab*, and *gtf3aa;gtf3ab* larvae at 5 dpf, showing developmental abnormalities in *gtf3aa* and *gtf3aa;gtf3ab* crispants. (**D**) Standard body length of larvae at 5 dpf. (**E**) Kaplan-Meier survival analysis of the indicated genotypes. (**F**) Representative merged bright-field and EGFP fluorescence images, and corresponding EGFP-only images, of the thymic region at 5 dpf demonstrating rescue of thymic EGFP expression in *gtf3aa* crispants following co-injection of 400 pg wild-type human *GTF3A* (hGTF3A) mRNA. (**G**) Quantification of the EGFP-positive area within the thymic region in rescued zebrafish larvae with the indicated control and crispant groups. Data in (A to F) are representative of at least two independent experiments. Mean ± SD of minimum 17 fish is shown. Statistical tests: Kruskall-Wallis test with Dunn’s multiple comparisons (D) and One-way ANOVA (G) with Dunnett’s multiple comparisons (B). ns: not significant, **P<0.01, ***P < 0.001, ****P<0.0001.

In addition to defective early thymocyte development, both *gtf3aa* and *gtf3aa;gtf3ab* crispants displayed marked developmental abnormalities, including reduced body length, uninflated swim bladder, and pericardial and yolk edema at 5 dpf (Fig. 5C, D). Scoring of the gross morphological phenotype demonstrated severe developmental defects in approximately 70% of *gtf3aa* and 75% of *gtf3aa;gtf3ab* crispants (Supplementary Fig. 6D), whereas *gtf3ab* crispants developed normally without apparent morphological abnormalities (Fig. 5C, D). These defects persisted at 8 dpf (Supplementary Fig. 6E). To further characterize the observed developmental defects, we performed Alcian blue staining at 8 dpf to visualize cartilage structures. This revealed severe craniofacial malformations in *gtf3aa* crispants, affecting Meckel’s cartilage, the palatoquadrate, ceratohyal, and ceratobranchials (Supplementary Fig. 6F). In line with these developmental abnormalities, both *gtf3aa* and *gtf3aa;gtf3ab* crispants displayed increased lethality after 5 dpf, whereas survival of *gtf3ab* crispants was comparable to controls (Fig. 5E).

To confirm that the reduced EGFP signal in the thymic region and the observed developmental abnormalities were specifically due to the loss of *gtf3aa,* and to exclude off-target effects, we performed rescue experiments by supplementing *gtf3aa* crispants with human *GTF3A* mRNA. Expression of human TFIIIA restored the thymic EGFP signal to near-control levels (Fig. 5F, G) and similarly rescued the developmental abnormalities observed in *gtf3aa* crispants (Supplementary Fig. 7A, B).

Altogether, our *in vivo* data in zebrafish confirm that *gtf3aa* is required for effective early T cell development and demonstrate that loss of *gtf3aa* disrupts normal cartilage skeletal structures development. The rescue of the early thymocyte development by complementing the zebrafish larvae with human *GTF3A* underscores the functional conservation of TFIIIA across species and establishes a conserved, non-redundant role for TFIIIA in early T cell development in the thymus.

## DISCUSSION

TFIIIA has long been recognized as the dedicated transcription factor and chaperone for 5S rRNA synthesis and assembly (*11–13*). Although recent work has implicated TFIIIA in innate immunity signaling (*21*, *22*), its potential role in adaptive immune development has remained unknown. Here, we identify biallelic rare *GTF3A* variants as a novel genetic cause of SCID and CID, uncovering a previously unrecognized role for TFIIIA in adaptive immunity. Across all patients, TFIIIA deficiency manifested as a consistent T cell defect, characterized by profound CD4⁺ T cell lymphopenia, variable CD8⁺ T cell involvement, markedly reduced naïve CD4^+^ and CD8^+^ T cells, diminished CD31⁺ RTE, low to absent TREC levels, and a restricted TCR repertoire.

Consistent with this phenotype, ATO cultures revealed a severe early developmental block at the stage of full T cell commitment, marked by persistent failure of CD1a upregulation. This is compatible with an inability of patient-derived progenitors to advance beyond early double negative thymocyte stages. Concordantly, *gtf3aa*-deficient zebrafish crispants expressing EGFP under control of the *lck* promotor exhibited a profound impairment in early thymocyte development, a defect fully rescued by expression of human *GTF3A*. In zebrafish, *lck* expression marks thymocytes shortly after thymus colonization, and murine studies have more precisely localized proximal *lck* promoter activity to early double-negative stages during progression towards T lineage commitment (*41*, *44*, *45*). Similarly, CD1a upregulation in human thymocytes marks the transition from multipotent progenitors to T lineage-committed cells (*46*). Supporting these findings, publicly available single-cell transcriptomic datasets of human fetal thymus development (cf. hosted on the UCSC Cell Browser (*47*)) show high *GTF3A* expression in early thymocyte populations, particularly within double-negative stages (Supplementary Fig. 8) (*48*). When positioned relative to other monogenic SCID disorders, TFIIIA deficiency appears to disrupt thymocyte development upstream of recombination defects such as RAG1/2 deficiency or metabolic defects such as Adenosine Deaminase (ADA) deficiency, which impair development at or beyond the double-positive stage, but downstream of signaling defects such as IL2RG deficiency, which arrest development at the level of T cell-specified progenitors (*40*). Together, these findings identify an essential role for TFIIIA in early human T cell ontogeny.

In contrast to the uniform T cell lymphopenia, the B cell phenotype was more heterogeneous across SCID and CID patients. B cell numbers were largely preserved in SCID patients, consistent with T^-^B^+^NK^+^ SCID, whereas they were reduced in CID patients, suggesting that B cell vulnerability emerges secondarily to the primary T cell defect and becomes more apparent with age. It is important to note, however, that definitive conclusions regarding later-onset B cell involvement in SCID patients were limited due to early HSCT or mortality. Bone marrow analysis in a limited number of patients revealed no evidence for impaired early B cell development, and peripheral immunophenotyping demonstrated increased transitional B cells, arguing against a major developmental block. Nevertheless, all evaluated CID patients exhibited impaired vaccine-specific antibody responses, particularly against polysaccharide antigens, indicating clinically relevant humoral dysfunction despite relatively preserved peripheral B cell development. This functional antibody defect may partly reflect defects of marginal zone B cells (not investigated here) and/or impaired T cell help, as suggested by the consistent reduction in circulating T follicular helper cells. However, the consistent IgM deficiency across patients suggests a degree of intrinsic B cell susceptibility that warrants further investigation.

Beyond the immune phenotype, *gtf3aa*-deficient zebrafish crispants displayed a broad spectrum of developmental abnormalities, including severe cartilage defects, and did not survive beyond early larval stages. These findings align with recent work by Luo *et al*., who reported that stable *gtf3aa*^⁻/⁻^ zebrafish exhibit severe multi-organ developmental abnormalities and early lethality (*49*). In a similar vein, attempts to generate complete *Gtf3a* knockout mice resulted in early embryonic lethality (data not shown), collectively indicating that complete loss of TFIIIA is incompatible with normal vertebrate development (*50–52*). The genetic architecture of our patient cohort supports this notion. Although patient-derived variants impair TFIIIA function, complete loss of TFIIIA activity has not been observed in a homozygous state to date: truncating variants occur only in *trans* with missense alleles, suggesting that full deficiency is not tolerated in humans either. This positions TFIIIA within a dosage-sensitive functional window in which residual activity is sufficient to support general organism development but is insufficient to sustain T cell development. In this light, the isolated non-immune features observed in our cohort may represent subtle manifestations of partial TFIIIA dysfunction, though no consistent syndromic features were identified across the cohort and P5’s short stature is more plausibly attributable to chronic illness. Whether additional features emerge more prominently in future patients harboring more hypomorphic alleles remains an open question that careful longitudinal phenotyping will need to address.

Mechanistically, the identification of 14 distinct disease-associated *GTF3A* variants distributed across zinc-finger domains, linker regions, and the C-terminal domain provided a unique opportunity to dissect TFIIIA function in a human disease context. Functional analysis revealed defects in several core TFIIIA activities, including (i) impaired DNA binding, (ii) reduced 5S rRNA binding and chaperone function, (iii) compromised protein stability, and (iv) defective nuclear import. Importantly, these functional consequences closely mirrored domain-specific roles previously inferred from model organisms (*12*, *16*, *18–20*), highlighting the evolutionary conservation of TFIIIA architecture and, for the first time, validating these mechanistic principles directly in human cells.

Despite this mechanistic diversity at the protein level, pathogenic *GTF3A* variants converge on a shared molecular outcome: reduced transcriptional output of canonical 5S rRNA and its paralog, *RNA5SP141*. Given the unique and non-redundant role of TFIIIA in 5S rRNA transcription, this shared defect likely represents the central pathogenic axis of TFIIIA deficiency. For many variants, impaired transcription could be readily explained by disruptions in core TFIIIA functions such as DNA binding or nuclear import. However, some variants – most notably the recurrent substitutions affecting residue R319 – consistently reduced transcriptional output despite preserved activity across functional assays. These findings suggest the existence of additional or more subtle mechanisms of TFIIIA dysfunction, potentially involving impaired assembly or stability of the RNA polymerase III transcriptional machinery rather than disruption of DNA binding or nuclear localization.

Although the transcriptional defect extends beyond canonical 5S rRNA to include *RNA5SP141,* an endogenous RIG-I ligand implicated in antiviral innate immune signaling (*21*, *22*), overt clinical manifestations of impaired innate antiviral defense were uncommon in our patient cohort. This apparent incomplete penetrance, although frequently observed in monogenic defects of antiviral immunity (*53*, *54*), may partly reflect limited clinical ascertainment in patients with SCID, who underwent early HSCT following newborn screening or died in early infancy, thereby restricting viral exposure and limiting the opportunity to clinically manifest defects in antiviral defense.

Given the canonical role of 5S rRNA as a core structural component of the 60S large ribosomal subunit and its essential function in ribosome biogenesis (*10*), TFIIIA deficiency can be situated within the broader landscape of ribosomopathies. Indeed, Luo *et al*. demonstrated that *gtf3aa^⁻/⁻^* zebrafish exhibit reduced 5S rRNA transcription accompanied by marked defects in ribosome assembly, including decreased levels of the 60S large subunit, mature 80S ribosomes, and actively translating polysomes (*49*). Yet, in contrast to well-studied ribosomopathies such as Diamond-Blackfan anemia or Shwachman-Diamond syndrome which typically present with multisystem syndromic features, TFIIIA deficient patients display a striking immune-selective phenotype (*33*, *55–57*). This discrepancy suggests that developing thymocytes may be uniquely sensitive to TFIIIA dysfunction, an interpretation supported by the increased *GTF3A* expression observed in early thymocyte populations potentially reflecting their high demands for ribosome biogenesis during rapid proliferation and differentiation (*48*). Alternatively, non-canonical functions of TFIIIA and/or 5S rRNA, involving pseudogene-derived transcripts or nucleolar stress signaling pathways, may further shape disease expression (*22*, *58*). Together, these observations position TFIIIA deficiency at an underexplored interface between ribosome biology and adaptive immune development, underscoring the need for future mechanistic studies to define what confers this immune selectivity.

In conclusion, we establish *GTF3A* as a novel monogenic cause of SCID and CID, and define an essential, previously unrecognized, role for TFIIIA in early human T cell development. By delineating TFIIIA dysfunction and revealing an immune-selective phenotype, this work expands the genetic and mechanistic landscape of inborn errors of immunity. From a clinical perspective, TFIIIA deficiency should be considered across a phenotypic spectrum of immunodeficiency, ranging from infants detected by TREC-based newborn screening who present with a T^-^B^+^NK^+^ SCID and no overt syndromic features, to children and adolescents with CID characterized by both T and B cell defects, severe infections, and immune dysregulation. Future studies will be required to define the precise mechanisms by which perturbations in TFIIIA function selectively compromise T cell development.

## MATERIAL AND METHODS

### Study approval

This study received ethical approval from the Ethics Committee of Ghent University Hospital (Ghent, Belgium; BC-11792), the institutional ethics committees of Necker Hospital for Children (Paris, France), and The Rockefeller University Hospital (New York, USA). Clinical data and patient samples were collected with informed consent from all participants in their countries of residence, in accordance with the Declaration of Helsinki (1975), local regulations, and NIH IRB-approved protocol (NCT03394053). All zebrafish experiments were conducted in compliance with EU Directive 2010/63/EU and were approved by the Ghent University Animal Ethics Committee (ECD 25-10K).

### Sequencing

Whole-exome sequencing (WES) or whole-genome sequencing (WGS) was performed in participating centers using standard protocols. Candidate variants in *GTF3A* (NM_002097.3) were validated by Sanger sequencing. Segregation analyses were conducted in available family members to confirm variant inheritance and consistency with the observed phenotype.

### Cell culture

Primary human dermal fibroblasts were cultured in Dulbecco’s modified Eagle’s medium (DMEM) supplemented with 10% fetal calf serum (FCS), 1% penicillin-streptomycin (10,000 U/ml, Gibco; 15140122), 2 mM GlutaMAX, 0.1 mM sodium pyruvate (Gibco; 11360070), and 50 µM 2-mercaptoethanol (Gibco; 31350010). HEK293T cells were maintained in DMEM supplemented with 10% FCS, 1% penicillin-streptomycin (10,000 U/ml), and 2 mM GlutaMAX (Gibco). TFIIIA C195W/C219R double knock-in (KI) HEK293T cells were previously described by Naesens et al. (*21*). MS5-hDLL4 (*59*) and MS5-hDLL1 (Merck; SCC167) stromal cells were cultured in MEMα medium (Gibco; 22561-021) containing 10% FCS and 1% penicillin-streptomycin (10,000 U/ml). All cell lines were cultured at 37°C and 5% CO2.

### Plasmids and cloning

The cDNA sequence encoding wild-type (WT) TFIIIA was cloned into a modified pcDNA3.1(+) expression plasmid containing an N-terminal GFP-tag (GenScript). Plasmids were propagated in *Escherichia coli* DH5α and purified using the NucleoSpin Plasmid EasyPure kit (Macherey-Nagel). TFIIIA variants were introduced into the WT TFIIIA plasmid using the Q5 Site-Directed Mutagenesis Kit, following the manufacturer’s instructions (New England Biolabs (NEB), E0552S). DNA concentration and purity were determined spectrophotometrically using the NanoDrop 2000 (Nanodrop Technologies). HEK293T cells were transiently transfected with either WT or mutant TFIIIA constructs using Lipofectamine 2000 (Invitrogen).

### RT-qPCR analysis

*RNA isolation and RT-qPCR.* Cells were lysed in RLT buffer (Qiagen) and stored at -80°C until further processing. Total RNA, including all small RNAs, were isolated using the miRNeasy Tissue/Cells Advanced Kit (Qiagen; 217684), according to the manufacturer’s instructions. RNA concentration and purity were assessed using a NanoDrop 2000 spectrophotometer. Equal amounts of RNA were reverse-transcribed into cDNA using the SensiFast cDNA Synthesis Kit (Bioline; BIO-65054) and quantitative real-time PCR (qPCR) was performed using a LightCycler 480 (Roche). Gene transcript levels were normalized to the average of housekeeping genes *HPRT*, *GAPDH*, and *β-actin*. All primers were purchased from Integrated DNA Technologies (IDT) and are listed in Supplementary Table 4.

*EU pulse chase.* HEK293T cells and primary fibroblasts were incubated with 0.5 mM 5-ethynyl uridine (EU) for 1 hour to label nascent RNA transcripts prior to RNA isolation. Total RNA was isolated using the miRNeasy Tissue/Cells Advanced Kits (217684; Qiagen) following the manufacturer’s instructions. Using the Click-iT Nascent RNA Capture Kit (C10365; Invitrogen), EU-labeled RNA was biotinylated via click chemistry and subsequently captured using streptavidin-coated magnetic beads. Isolated EU-labeled RNA was reverse-transcribed into cDNA using the SensiFast cDNA Synthesis Kit and qPCR was performed on a LightCycler 480. Gene transcript levels were normalized to the average of housekeeping genes *HPRT*, *GAPDH*, and *β-actin*.

*RNA5SP141 expression.* RNA was isolated using miRNeasy Tissue/Cells Advanced Kits. Reverse transcription reactions were done using *RNA5SP141* and *Let-7a* specific TaqMan MicroRNA Assay primers, followed by RT-qPCR analysis using *RNA5SP141* (ThermoFisher cat #CSN1ESE) and *Let-7a* (ThermoFisher cat #4440887) specific TaqMan FAM reporter dye probes and TaqMan Fast Advanced Master Mix. Fold expression levels of *RNA5SP141* were normalized to *Let-7a* expression.

### EMSA

HEK293T cells were seeded at a density of approximately 78,000 cells/cm² and forward transfected with the indicated pcDNA3.1(+) plasmids using Lipofectamine 2000 (Invitrogen). After 16 hours, cells were lysed in radioimmunoprecipitation assay (RIPA) buffer (150 mM NaCl, 1.0% IGEPAL CA-630, 0.5% sodium deoxycholate, 0.1% SDS, and 50 mM Tris, pH 8.0) (Roche) supplemented with protease inhibitors (cOmplete ULTRA; Roche). Protein lysates (10 µg) were incubated with an IRDye 700-labeled 5S rRNA oligonucleotide (5′-AAGCTAAGCAGGGTCGGGCCTGGTTAGTACTTGGATGG-GAGACCGC-3′) in binding buffer (100 mM Tris, 500 mM KCl, and 10 mM dithiothreitol (DTT; pH 7.5)) supplemented with 25 mM DTT/2.5% Tween 20, MgCl_2_, NP-40, and poly(I:C) (1 μg/ml). Following a 30 min incubation at room temperature, Orange Loading Dye was added, the samples were loaded onto a 6% polyacrylamide gel and electrophoresis (MiniPROTEAN, Bio-Rad) was performed at 100 V for 90 min. Signal detection and quantification were performed using the Odyssey infrared imaging system (LI-COR). Equal protein input for WT and mutant TFIIIA was confirmed by immunoblotting. All EMSA experiments were repeated at least twice, and representative results are shown.

### Immunoblotting

Cells were lysed in RIPA buffer supplemented with protease inhibitors (cOmplete ULTRA; Roche) and phosphatase inhibitors (PhosSTOP; Roche). The soluble protein fraction was collected and normalized prior to electrophoretic separation. Samples were resolved by SDS-PAGE on 4-15% Criterion TGX Stain-Free Protein Gels (Bio-Rad) and subsequently transferred to nitrocellulose membranes using a semi-dry transfer system (Bio-Rad). Membranes were blocked for 1 hour at room temperature in phosphate-buffered saline (PBS) containing 0.1% Tween 20 and 10% nonfat dry milk (Cell Signaling Technology (CST)). Primary antibody incubation was performed either overnight at 4°C or for 2 hours at room temperature, followed by incubation with horseradish peroxidase (HRP)-conjugated secondary antibodies. Membranes were developed using enhanced chemiluminescence (ECL) substrates (SuperSignal West Dura or Femto; Thermo Fisher Scientific) and visualized on a Chemidoc imaging system (Bio-Rad). All immunoblotting experiments were independently repeated at least twice, and representative data are presented. All uncropped images can be retrieved in the Source File. The following primary antibodies were probed: anti-TFIIIA (1:1,000; ab129440; Abcam), anti-GFP (1:500; MA5-15256; Invitrogen), and anti-GAPDH-HRP (1:2,000; #8884; CST). The secondary antibodies goat anti-mouse IgG (H+L)-HRP (1:10,000; G-21040; Invitrogen) and goat anti-rabbit IgG-HRP (1:2,000; #7074; CST) were used.

### RNA pull-down

RNA-protein interaction assays were performed using the Pierce^TM^ Magnetic RNA–Protein Pull-Down Kit (#20164, Thermo Fisher Scientific), following the manufacturer’s protocol. A custom 5′-biotinylated 5S rRNA oligonucleotide (IDT) was used. For each assay, 50 pmol of RNA was pre-bound to 50 μL of streptavidin magnetic beads prior to incubation with cell lysates. HEK293T cell lysates were prepared in RIPA buffer supplemented with protease (cOmplete ULTRA; Roche) and phosphatase inhibitors (PhosSTOP; Roche). 150 μg of total protein was incubated with RNA-coated beads under RNase-free conditions. After washing to remove non-specific interactions, bound proteins were eluted in 4x Laemmli Sample Buffer (Bio-rad) and analyzed by immunoblotting.

### Confocal imaging

HEK293T cells were seeded at a density of approximately 60,000 cells/cm² in eight-well chamber slides (Ibidi) and forward-transfected after 24 hours with the indicated pcDNA3.1(+) plasmids using Lipofectamine 2000 (Invitrogen). After 16 hours, cells were fixed with 3% paraformaldehyde (PFA) in PBS for 20 min, followed by permeabilization in 0.2% Triton X-100 for 10 min and blocking in 0.5% BSA (Sigma-Aldrich) and 0.02% Triton X-100 in PBS for 30 min. Primary antibody incubation was performed overnight at 4°C with anti-NPM1 (1:250; CST; 92825). After washing, cells were incubated for 2 h at room temperature with secondary antibody goat anti-rabbit Alexa Fluor 568 (1:1000; Invitrogen; A-11036). Following washing, cells were stained with Hoechst 33342 nuclear stain (1:1000; Invitrogen) for 15 min at room temperature and subsequently mounted in PVA-DABCO (Sigma-Aldrich). Confocal images were acquired with a Zeiss LSM 980 Airyscan confocal microscope using a Plan-Apochromat 63×/1.4 oil objective. Z stacks were taken with an interval of 0.14 µm in Airyscan mode with a zoom of 1.7. Images were processed using Zen Zeiss software. Microscope settings and image processing steps were kept consistent for all samples of the same experiment.

### Deep immunophenotyping

*CyTOF*. Fresh heparin whole blood from two TFIIIA-deficient patients (P2, P3; obtained pre-HSCT) and age-matched healthy controls was used. All samples were processed within 24 hours of collection. Cells were stained using a custom-designed antibody panel as listed in Supplementary Table 5, according to the manufacturer’s instructions (Fluidigm). Labelled cells were frozen at -80°C after overnight live-dead staining. Acquisition was performed on a Helios mass cytometer (Fluidigm) and data was analyzed using OMIQ software. Gating for CyTOF immunophenotyping was performed as previously described (*60*).

*Flow cytometry*. Cryopreserved peripheral blood mononuclear cells (PBMCs) from three TFIIIA-deficient patients (P6, P7, P8) were thawed in preheated complete RPMI-1640 medium supplemented with GlutaMAX (Gibco; 61870-010), 10% FCS, 1% penicillin-streptomycin (10,000 U/mL), 1 mM sodium pyruvate, 1% non-essential amino acids (NEAA; Gibco; 11140035), and 50 μM 2-mercaptoethanol. Following thawing, cells were left to recover for 30 min at 37°C and 5% CO_2_. Subsequently, cells were counted and 2×10^6^ cells were used for staining. Cells were first incubated with FcR blocking reagent (BioLegend; 422302) in PBS, together with a biotin-conjugated antibody and Zombie UV™ Fixable Viability dye (BioLegend; 423107). Next, a first set of surface markers was stained with a mixture of antibodies in PBS containing Brilliant Stain buffer (BD Biosciences) for 30 min at 4°C. After washing, a second set of surface markers was stained overnight at 4°C. Lastly, cells were fixed and permeabilized, and intracellularly stained with antibodies using the Foxp3/Transcription Factor Staining Buffer Set (Invitrogen; 00-5523-00) according to the manufacturer’s protocol. Data acquisition was performed with a FACSymphony flow cytometer (BD biosciences) and data were analyzed with the FlowJo v10.10.0 software (BD Life Sciences). A detailed list of antibodies used to define PBMC populations is provided in Supplementary Table 6.

### T cell receptor (TCR) repertoire analysis

*TCR sequencing.* TCR sequencing was performed on three patients (P6, P7, P8) and six healthy controls, with analyses performed at the subject level. TCR libraries were generated following the TIRTL workflow and sequenced on a short-read, paired-end sequencing platform (*61*). Raw sequencing reads were processed using MiXCR to identify productive TCR clonotypes, defined by CDR3 amino acid sequence with assigned V and J gene usage. Analyses were performed separately for the TRA and TRB chains. For downstream analyses, wells originating from the same donor were pooled, yielding donor-level clonotype frequency distributions for each chain (TRA and TRB). From donor-level clonotype frequencies, repertoire metrics were calculated, including number of unique clonotypes (clonotype richness), Shannon entropy (clonotype diversity), and cumulative frequency of the top 1 and top 10 clonotypes (oligoclonality). Productive read depths were assessed to ensure comparability between samples and experimental groups prior to repertoire analysis.

#### TCR Vβ repertoire analysis

The T cell receptor Vβ (TCR-Vβ) repertoire was analyzed by flow cytometry using the IOTest β Mark TCR Repertoire Kit (IM3497, Beckman Coulter). PBMCs from patient P5 and healthy donors were stained with the TCR-Vβ antibody panel according to the manufacturer’s instructions. Cells were co-stained with anti-human CD45 V500 (clone HI30; BD Biosciences), CD3 BV421 (clone UCHT1; BD Biosciences), CD4 Alexa Fluor 700 (clone OKT4; eBioscience), and CD8α PE-Dazzle (clone RPA-T8; BioLegend) to identify TCR-Vβ family usage within CD45⁺CD3⁺CD4⁺ and CD45⁺CD3⁺CD8⁺ T cell populations. Repertoire skewing was quantified using the Gini-TCR skewing index as previously described (*62*).

#### Artificial thymic organoids

##### Isolation of hematopoietic progenitor cells

CD34^+^ hematopoietic stem and progenitor cells (HSPCs) were enriched from cryopreserved patient-derived PBMCs (P2, P6, P7, P8) or bone marrow mononuclear cells (BM MNCs; P5). Depending on the experiment, enrichment was performed either by fluorescence-activated cell sorting (FACS) or by magnetic separation using CD34 selection kits. For FACS-based enrichment, CD34^+^Lin^-^ cells were sorted on a BD FACSAria Fusion (BD biosciences), with lineage-negative (Lin⁻) being defined as negative for CD3, CD14, CD19, and CD56 expression. Magnetic enrichment was performed using either the EasySep Human Cord Blood CD34 Positive Selection Kit (Stemcell Technologies; 17896) or the CD34 MicroBead Kit UltraPure (Miltenyi Biotec) on an AutoMACS Pro Separator (Miltenyi Biotec), according to the manufacturer’s instructions. CD34⁺ cells from TFIIIA-deficient patients (PBMC-derived or BM MNCs) and healthy donors (PBMC-derived or mobilized peripheral blood) were used for Artificial thymic organoids (ATO) generation.

##### ATO assembly and culture

ATOs were assembled and cultured as previously described with minor modifications (*39*, *63*). MS5 stromal cells expressing human DLL1 (MS5-hDLL1) or DLL4 (MS5-hDLL4) were used. Briefly, 120-3,417 CD34^+^ cells were aggregated with 150,000 stromal cells per organoid. To promote T cell development, ATOs were cultured in either RPMI-1640 (Gibco), containing 4% B-27, 1% penicillin-streptomycin (10,000 U/ml), 1% GlutaMAX, L-ascorbic acid 2-phosphate (30 µM), IL-7 (5 ng/ml), and FLT3-L (5 ng/ml), or RB27 medium supplemented with L-ascorbic acid 2-phosphate (30 µM), IL-7 (5 ng/ml), and FLT3-L (5 ng/ml). Organoids were maintained for approximately 5-6 weeks before analysis.

##### Flow cytometric analysis

At defined time points, ATOs were harvested by mechanical dissociation using vigorous pipetting or by adding MACS buffer (PBS containing 0.5% BSA and 2 mM EDTA) followed by pipetting to dissociate the aggregates. For immunophenotyping of developing human T cell progenitors, cells recovered from ATOs were stained in Brilliant Stain buffer for with antibodies against CD45, CD3, CD4, CD8β, CD7, CD1a, and TCRαβ, together with lineage markers CD3, CD14, CD19, CD56, and CD34. Subsequently, dead cells were stained using propidium iodide (PI). Dead cells were excluded using either propidium iodide (PI) or LIVE/DEAD™ Fixable Yellow Dead Cell Stain (Invitrogen). Data were acquired on a BD LSR II Fortessa or BD FACSymphony flow cytometer (BD Biosciences) and analyzed using FlowJo software (BD Life Sciences). A complete list of antibodies used is provided in Supplementary Table 7.

##### Bone marrow analysis

Bone marrow samples from two TFIIIA-deficient patients (P3, P8) were analyzed by flow cytometry to assess B cell developmental stages. Data were interpreted according to the classification and gating strategy described by Wentick *et al*. (*64*).

#### Zebrafish studies

##### Zebrafish maintenance

Zebrafish were maintained in accordance with the Animal Research Guidelines at the Zebrafish Facility Ghent (ZFG), Ghent University (*65*, *66*). Zebrafish larvae were placed in E3 medium in an incubator at 28 °C until 5 days post fertilization (5dpf). After 5 days, they were housed in a semi-closed recirculating system (ZebTec, Tecniplast, Buguggiatte, Italy) at a constant temperature (27–28 °C), pH (∼7.5), and conductivity (∼550 mS), and kept on a standard light/dark cycle (14 h/10 h). The *Tg(lck:EGFP)* transgenic zebrafish line used in this study was previously described (*41*). All experiments were approved by the Animal Ethics Committee of the Ghent University Faculty of Medicine and Health Sciences (ECD 25-10K) and conform to the guidelines from Directive 2010/63/EU of the European Parliament on the protection of animals used for scientific purposes. Pain, distress, and discomfort were minimized as much as possible.

##### Generation of zebrafish crispants

CRISPR/Cas9-mediated genome editing was performed to generate *gtf3aa*, *gtf3ab*, and *gtf3aa*;*gtf3ab* zebrafish crispants. CRISPR guide RNAs (crRNAs) targeting the *gtf3aa* and *gtf3ab* genes were designed using the CRISPR Guide RNA Design Tool (Benchling) and the Alt-R™ CRISPR HDR Design Tool (IDT) using GRCz11 as a reference genome. A non-specific crRNA referred to as the ‘scrambled’ was used to generate the controls (*67*). The crRNAs and primers sequences used are listed in Supplementary Table 8 and 9. gRNA duplexes were formed by mixing each crRNA with trans-activating CRISPR-RNA tracrRNA (IDT), heated at 95 °C for 5 min, then cooling to room temperature. These duplexes (5 µM) were micro-injected with Cas9 protein (800 ng/µL; IDT) into one-cell stage Tg(*lck:EGFP*) zebrafish embryos, to deliver a total of 5 μM Cas9 protein with an equimolar amount of guide RNA duplex. To assess genome editing efficiency, genomic DNA was extracted from 10 pooled embryos at 1 dpf by denaturation in 50 mM NaOH at 95 °C for 20 min, followed by the addition of 10 ml 1 M Tris-HCl (pH 8) to neutralize the solution. Indel mutations at the region of interest were detected by PCR amplification followed by Sanger sequencing.

##### Thymic region imaging and standard length measurement

For live imaging, 5 and 8 dpf zebrafish larvae with normal heartbeat and blood circulation were selected and were laterally positioned and embedded in 1% low-melting temperature SeaPlaque™ Agarose (Lonza) supplemented with 120 mg/L tricaine (Sigma-Aldrich). Thymic fluorescent signal was acquired using an Axio Observer Z.1 microscope equipped with a monochrome Axiocam 506 camera (Carl Zeiss Microscopy GmbH) and surface areas were quantified using Fiji software (version 2.16.0). Standard length (SL) was defined as the distance from the snout to the base of the tail (*68*). SL measurements were obtained using bright-field images taken with a Leica M165 FC Fluorescent Stereo Microscope (Leica Microsystems) and quantified using Fiji software (version 2.16.0).

##### Alcian blue staining

After live imaging, 5 and 8 dpf zebrafish were euthanized using 4 g/L Tricaine. Larvae were transferred to a cell strainer, rinsed thoroughly with dH_2_O to remove residual Tricaine, and fixed overnight in 4% paraformaldehyde (PFA) at room temperature with gentle agitation. Following washing with dH_2_0, the larvae were bleached for 20-25 min at room temperature in dH_2_O containing 30% H_2_O_2_ and 10% KOH. Subsequently, larvae were incubated overnight at 4 ℃ in Alcian blue staining solution (0.1% Alcian blue and 1% HCl in 70% ethanol). Following staining, larvae were rinsed for 3.5-4 h in de-staining solution (5% HCl in 70% ethanol), then sequentially washed in 50% ethanol/50% PBST, followed by 25% ethanol/75% PBST, each at room temperature. Final washes were performed with dH₂O prior to stepwise transfer into 50%, 70%, and 100% glycerol (5 min each). Imaging was performed using a Leica M165FC microscope (Leica Microsystems).

#### Zebrafish rescue

##### Plasmid and cloning

The coding sequence of human *GTF3A* was analyzed in silico for the presence of *BamHI* or *BspQI* restriction sites as well as possible terminator sequences. Where necessary, synonymous codon substitutions were introduced to remove these sites while preserving the amino acid sequence. The modified sequence was subsequently checked for proper RNA folding and custom-synthetized as gBlock (IDT). To allow monitoring of mRNA delivery *in vivo*, the gBlock was designed to include an N-terminal mCherry fluorescent reporter, fused in-frame to the *GTF3A* coding sequence via a short GS linker. Next, the gBlock was cloned into a modified pST1 mRNA vector backbone (a kind gift from Prof. K. Thielemans, VUB) using the Gibson Assembly method (NEB). Transformation into competent *Escherichia coli* DH10β cells was performed followed by plasmid DNA purification (Qiagen). Quality control included Sanger sequencing as well as gel electrophoresis after restriction digest. DNA concentration and purity were determined using the Nanodrop ND-1000 spectrophotometer (ThermoFisher).

##### In vitro transcription of mRNA

Plasmid DNA was linearized with the *BspQI* restriction enzyme (NEB) and used as a template for *in vitro* transcription using the Hiscribe T7 mRNA kit with CleanCap reagent AG (NEB; E2080S), following the manufacturer’s protocol. RNA concentration and purity were determined using the Nanodrop ND-1000 spectrophotometer and RNA integrity was confirmed using a Bioanalyzer 2100 (Agilent). Endotoxin levels were evaluated using the Endosafe ® kinetic chromogenic cartridge technology (Charles River Technologies).

##### Zebrafish mRNA complementation

Tg(*lck*:*EGFP*) zebrafish embryos were co-injected at the one-cell stage with *gtf3aa* gRNA duplex and Cas9 protein (concentrations as described above), together with 400 pg of *in vitro*-transcribed human *GTF3A* mRNA. Successful mRNA delivery was verified by monitoring mCherry fluorescence using a Leica M165 FC Fluorescent Stereo Microscope (Leica Microsystems) at 6, 24, 48, and 72 hours post-fertilization. At 5 dpf, embryos were imaged and data was quantified as described above to assess the effect of mRNA complementation.

## Statistical analyses

All statistical analyses were performed using GraphPad Prism software (v10). Specific statistical tests used are detailed in the corresponding figure legends. Results were considered statistically significant at p < 0.05, with significance levels indicated as follows: p < 0.05 (*), p < 0.01 (**), p < 0.001 (***), and p < 0.0001 (****). Representative western blots, flow cytometric plots, and immunofluorescence images are shown.

## Data Availability

All relevant data are available within the main text and supplementary materials. Additional datasets, materials, or protocols can be obtained from the corresponding authors upon reasonable request.

## Acknowledgments

We gratefully thank the patients and their families for participating in and consenting to this research. We thank Ludivine Wacheul (RNA Molecular Biology, ULB), Monique van Ostaijen-ten Dam (Leiden University Medical Center), Mélanie Migaud (Imagine institute), and Jeremy Bertrand (SeqOIA Genomics Laboratory) for technical assistance, and Sylvie De Buck (UGent), Karlien Claes (UGent), Tracey-Ann Höltermann (NIH), Yelena Nemirovska, Candace Clift, Lazaro Lorenzo-Diaz, Maya Chrabieh, Amyrath Geraldo, and Mark Woollett (Imagine Institute and Rockefeller University) for administrative support. We acknowledge the Core Zebrafish Facility Ghent, the Ghent University Flow Cytometry Core facility, the VIB Flow Core, VIB Bioimaging Core of the VIB-UGent Center for Inflammation Research, and the VVTG platform at INEM (Paris Cité University, France) for their assistance and expertise. This work was supported (non-financially) by the European Reference Network for Rare Immunodeficiency, Autoinflammatory, and Autoimmune Diseases Network (ERN-RITA).

## Funding

This work was supported by the European Research Council (ERC-101221534), the VIB Grand Challenge (VIB-UGent), CSL Behring, the GENeHOPE fund (King Baudouin Foundation), and in part by the Intramural Research Program of the National Institutes of Health (NIH). S.J.T. is a beneficiary of a postdoctoral FWO grant (1236923N). Centre for Primary Immune deficiency Ghent (CPIG) is recognized as Jeffrey Modell Diagnostic and Research center and funded by JMF foundation. The Laboratory of Human Genetics of Infectious Diseases is funded by the National Institute of Allergy and Infectious Diseases (R01AI095983), the National Center for Advancement of Translational Sciences (UL1TR001866), the Rockefeller University, the St. Giles Foundation, INSERM, Paris Cité University, the French National Research Agency (ANR) under France 2030 program (ANR-10-IAHU-01), and Laboratoire d’Excellence Integrative – Biology of Emerging Infectious Diseases (ANR-10-LABX-62-IBEID). T cell team Taghon is funded by the special research fund of Ghent University (BOF BAF and GOA) and the Fund for Scientific Research Flanders (FWO grant G0A1725N). The Antwerp Center for Translational Immunology and Virology is funded by FWO (G0AD025N). The contributions of the NIH authors are considered Works of the United States Government. The findings and conclusions presented in this paper are those of the author(s) and do not necessarily reflect the views of the NIH or the U.S. Department of Health and Human Services.

## Author contributions

Conceptualization: ED, SP, TJ, TT, JB, LDN, SJT, and FH Methodology: ED, SP, TJ, MB, BO, MVDB, SVL, KV, DLJL, TT, JB, LDN, SJT, and FH Investigation: ED, SP, MB, YVD, IV, VD, HL, SG, JB, SVL, LP, MW, HB, JR, and SJT Clinical data collection: ED, SP, RM, JR, NA, FP, LN, DB, OMD, IKC, SH, AH, MYK, AE, GG, CP, EJ, BN, AP, TK, DB, JLC, BL, JB, LDN, SJT, and FH Visualization: ED, SP, SJT, and FH Funding acquisition: ED, TT, JB, LDN, SJT, and FH Supervision: ED, TJ, PS, TT, JB, LDN, SJT, and FH Writing - original draft: ED, SP, SJT, and FH Writing - review & editing: All authors

## Competing interests

The authors declare that they have no competing interests.

## Data and materials availability

All *GTF3A* variants identified in this study will be submitted to ClinVar upon publication. All relevant data are available within the main text and supplementary materials. Additional datasets, materials, or protocols can be obtained from the corresponding authors upon reasonable request.

## Notes

### Competing Interest Statement

The authors have declared no competing interest.

### Author Declarations

The Ethics Committee of Ghent University Hospital, the Institutional Ethics Committee of Necker Hospital for Children, and the Institutional Ethics Committee of The Rockefeller University Hospital gave ethical approval for this work.

