## Supplementary Materials and Data for "Inherited human TFIIIA deficiency disrupts T cell development"

**The file includes:**

Supplementary Text
Supplementary Figures 1-8
Supplementary Tables 1-9

### SUPPLEMENTARY TEXT

#### Detailed case reports for patients with biallelic loss-of-function mutations in *GTF3A*

*Personally identifiable patient information was redacted in accordance with medRxiv requirements.*

### SUPPLEMENTARY FIGURES

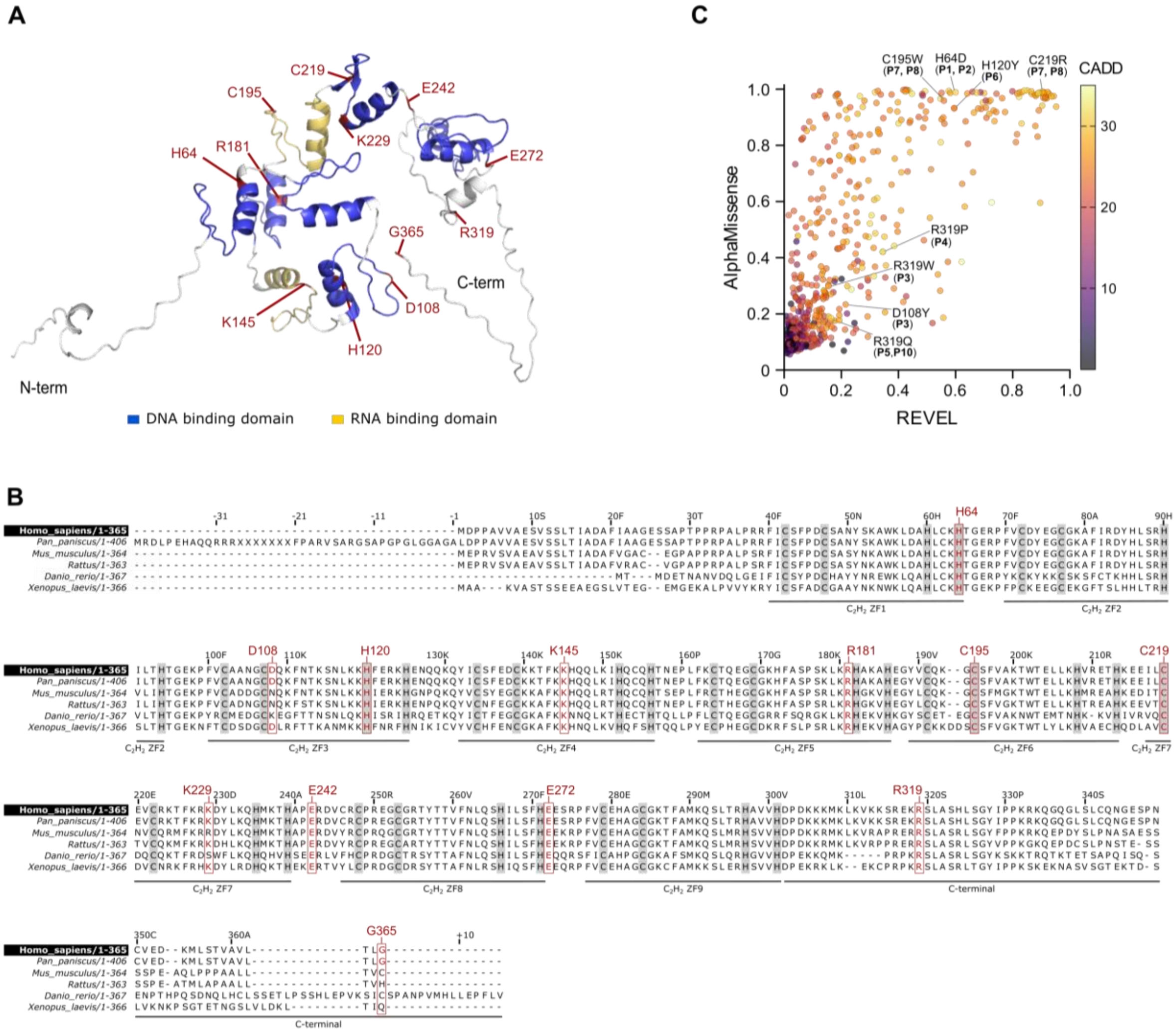


**Supplementary Figure 1 | Structural localization, evolutionary conservation, and *in silico* pathogenicity of *GTF3A* variants. (A)** Patient-identified *GTF3A* variants mapped onto the AlphaFold-predicted structure of the TFIIIA protein. Functional domains are indicated, with DNA-binding zinc-finger motifs shown in blue and RNA-binding zinc-finger motifs shown in yellow. (**B**) Multiple sequence alignment of TFIIIA orthologs from the indicated species generated using Clustal Omega, highlighting evolutionary conservation across zinc-finger domains. Conserved $C_2H_2$ residues shown in gray; *GTF3A* variants are indicated, with evolutionarily conserved amino acids shown in red. (**C**) *In silico* pathogenicity predictions for *GTF3A* missense variants identified in the patient cohort and those reported in gnomAD v4.1.0, with patient-derived variants indicated.

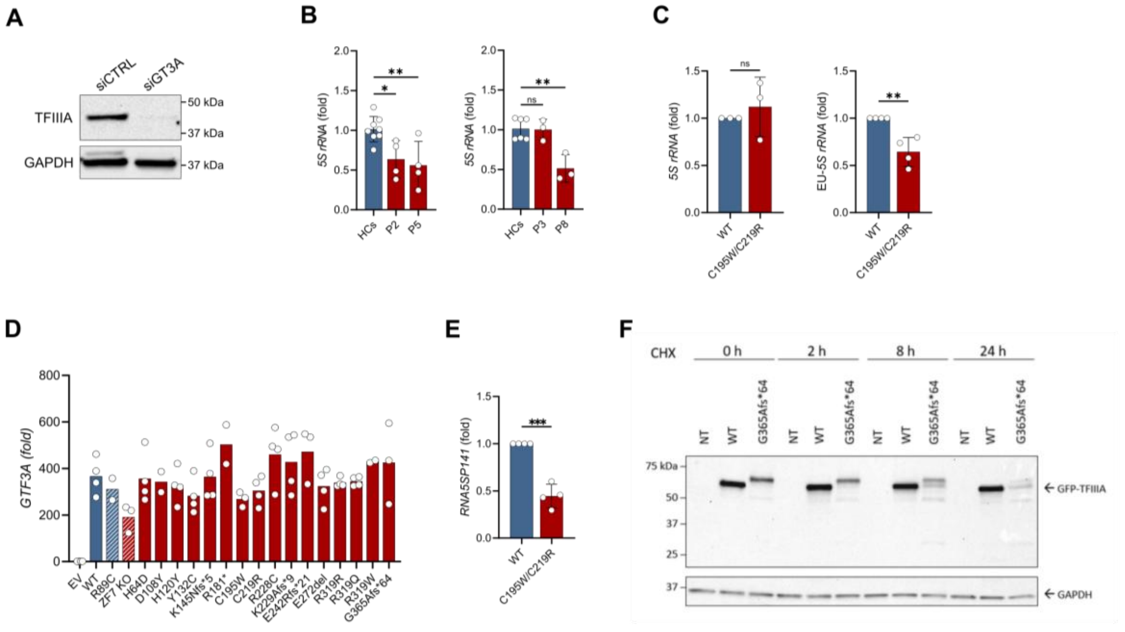


**Supplementary Figure 2 | Extended characterization of *GTF3A* variant effects on canonical 5S rRNA and *RNA5SP141* pseudogene transcription, and protein stability.** (**A**) Validation of TFIIIA antibody specificity by immunoblot in primary fibroblasts transfected with non-targeting control (siCTRL; 1 nM, 48h) or *GTF3A*-targeting siRNA (siGTF3A; 1 nM, 48h). GAPDH serves as a loading control. (**B**) RT-qPCR analysis of 5S rRNA expression in primary fibroblasts from patients (P2, P3, P5, and P8) compared to healthy controls (HCs, n=2). Pooled data from at least three independent repeats is shown. (**C**) RT-qPCR analysis of baseline (left) and nascent (right) 5S rRNA expression in WT and C195W/C219R knock-in (KI) HEK293T cells following EU labeling and pull-down. Pooled data from at least three independent repeats is shown. (**D**) RT-qPCR analysis of *GTF3A* expression in TFIIIA C195W/C219R KI HEK293T cells that were transfected for 16 hours with EV, GFP-tagged WT, or mutant TFIIIA constructs. Pooled data from at least two independent repeats per variant is shown. (**E**) RT-qPCR analysis of *RNA5SP141* expression in TFIIIA WT and C195W/C219R KI HEK293T cells. Pooled data from four independent repeats is shown. (**F**) Cycloheximide (CHX)-chase assay performed on WT HEK293T cells transfected with either GFP-tagged WT, the G365Afs*64 variant, or non-transfected (NT), demonstrating reduced mutant protein stability. Data are representative of two repeats. Statistical test: One-way ANOVA with Dunnett's multiple comparisons (A) and Student's t-test (B, D). ns: not significant, *P<0.05, **P<0.01, ***P<0.001.

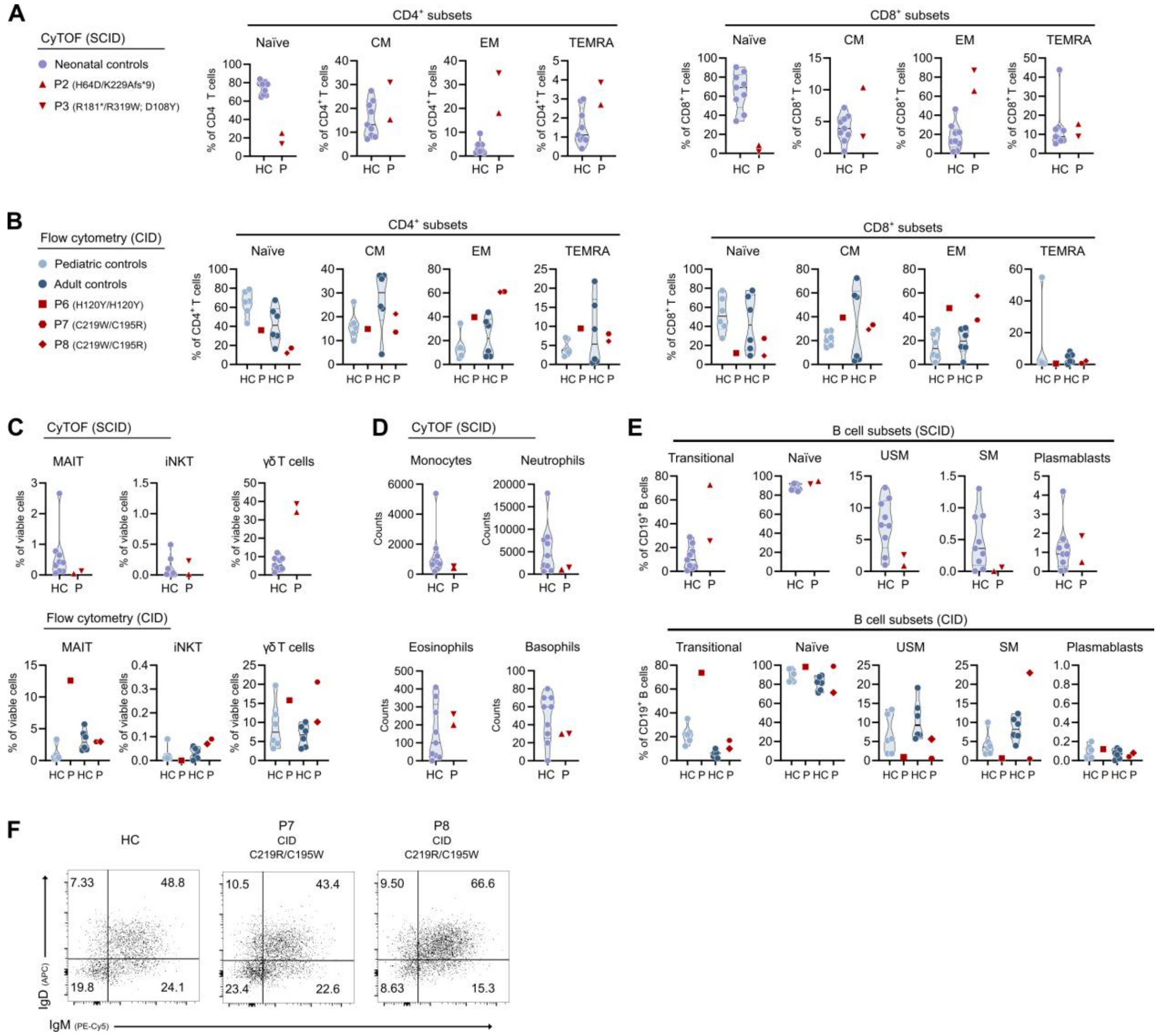


**Supplementary Figure 3 | In-depth immunophenotyping of TFIIIA-deficient SCID and CID patients.** (**A-C**) Frequencies of T cell subsets in whole blood or peripheral blood mononuclear cells (PBMCs) from two TFIIIA-deficient SCID patients (P2, P3) or three CID patients (P5, P6, P7), respectively. Age-matched neonatal (n=9), pediatric (n = 6), and adult controls (n = 6) were included. SCID patients were analyzed by Cytometry by Time of Flight (CyTOF); CID patients were analyzed by 27-parameter multiparametric flow cytometry with manual gating. T cell subsets were defined as naïve ($CCR7^{+}CD45RA^{+}$), central memory (CM; $CCR7^{+}CD45RA^{-}$), effector memory (EM; $CCR7^{-}CD45RA^{-}$), terminally differentiated effector memory (TEMRA; $CCR7^{-}CD45RA^{+}$), mucosal-associated invariant T cells (MAIT; $CD161^{+}V\alpha 7.2^{+}$), invariant natural killer T cells (iNKT; $CD161^{+}V\alpha 24J\alpha 18^{+}$), and γδ T cells ($CD3^{+}TCR\gamma\delta^{+}$). (**D**) Absolute counts of monocytes, neutrophils, eosinophils, and basophils in whole blood from two TFIIIA-deficient SCID patients (P2, P3) and age-matched neonatal controls (n=9), as determined by CyTOF. (**E**) B cell subset distribution in SCID, patients, CID patients, and age-matched controls, analyzed by either CyTOF or flow cytometry. B cell subsets were defined as transitional ($CD27^{-}CD24^{+}CD38^{+}$), naïve ($CD27^{-}CD24^{-}CD38^{-}$), unswitched memory (USM; $CD27^{+}IgD^{+}$), switched memory (SM; $CD27^{+}IgD^{-}$), and plasmablasts ($CD27^{-}IgD^{-}CD38^{+}$). Data (A-E) are representative of a single experiment. (**F**) IgM surface expression on resting $CD19^{+}$ B cells of two TFIIIA-deficient patients (P7 and P8) compared to an adult healthy control (HC).

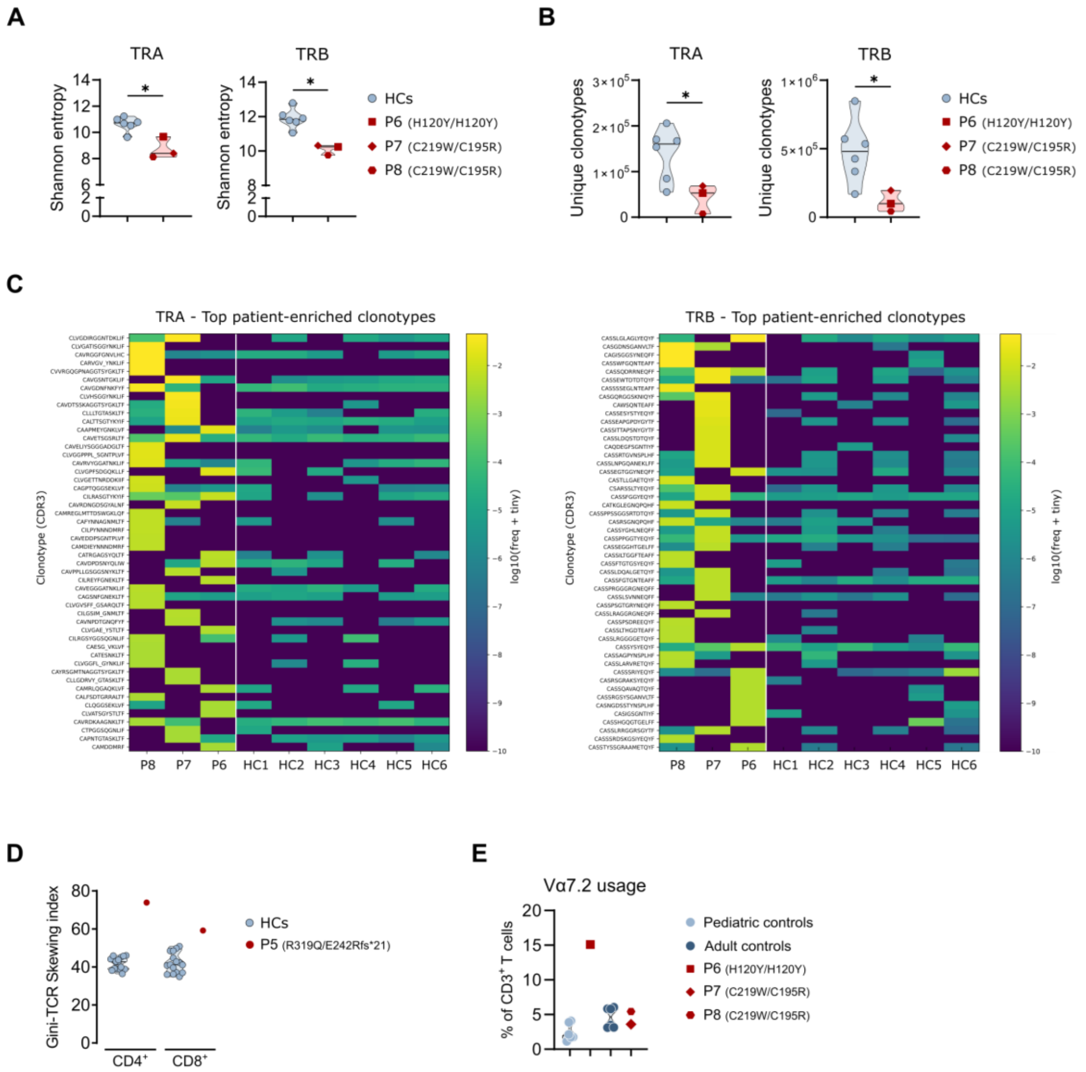


**Supplementary Figure 4 | Restricted TCR repertoire diversity and increased clonotype dominance in TFIIIA-deficient CID.** (**A**) Shannon entropy of productive TCR repertoires for the TRA and TRB chains, determined from peripheral blood T cells of TFIIIA-deficient CID patients (P6, P7, and P8) and age-matched HCs, demonstrating reduced repertoire diversity in patients. (**B**) Clonotype richness, defined as the number of unique productive clonotypes, for TRA and TRB repertoires in TFIIIA-deficient patients and HCs. (**C**) Heatmaps showing the relative frequencies of the top patient-enriched productive TCR clonotypes for the TRA (left) and TRB (right) chains. Clonotypes were ranked based on their cumulative frequency in patients, with $\log_2$ patient/control frequency ratio used. For each chain, the top 50 clonotypes were selected and visualized across individual donors. (**D**) Gini-TCR skewing index showing TRB V-gene usage in $CD4^+$ and $CD8^+$ T cell subsets from CID patient P5 and healthy controls. (**E**) Vα7.2 usage in three CID patients (P6, P7, P8) compared to age-matched pediatric and adult controls, analyzed by flow cytometry. Data (A-E) are representative of one experiment. Statistical test: Mann-Whitney U with Benjamini-Hochberg FDR correction (A, B). $^*P<0.05$.

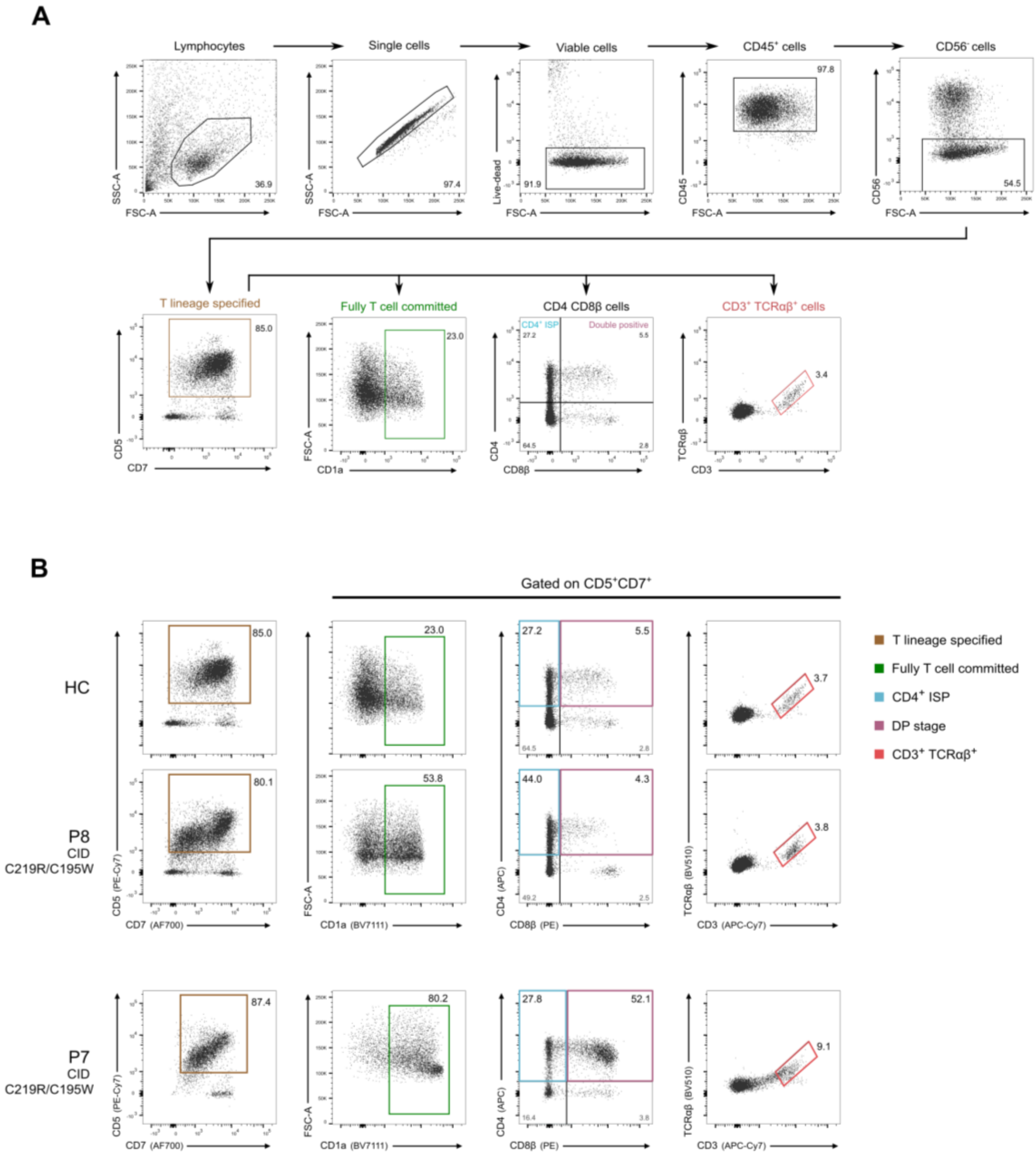


**Supplementary Figure 5 | Block in T cell development in TFIIIA-deficient human hematopoietic progenitors.** (**A**) Representative flow cytometry gating strategy for *in vitro* T cell differentiation of healthy control (HC) CD34+ progenitors cultured in the artificial thymic organoid (ATO) system. Cells were gated on LIVE/DEAD-CD45+CD56- events and assessed for expression of early and late T lineage commitment markers, including CD7, CD5, CD1a, CD4, CD8α, CD8β, CD3, and TCRαβ. (**B**) T cell differentiation of CD34+ progenitors from TFIIIA-deficient patients from family 6. Flow cytometry plots show expression of CD7, CD5, CD1a, CD4, CD8β, TCRαβ, and CD3 within LIVE/DEAD-CD45+CD56- cells (analyzed at 5 week). ISP: immature single positive (CD4+CD8β−); DP: double positive (CD4+CD8β+).

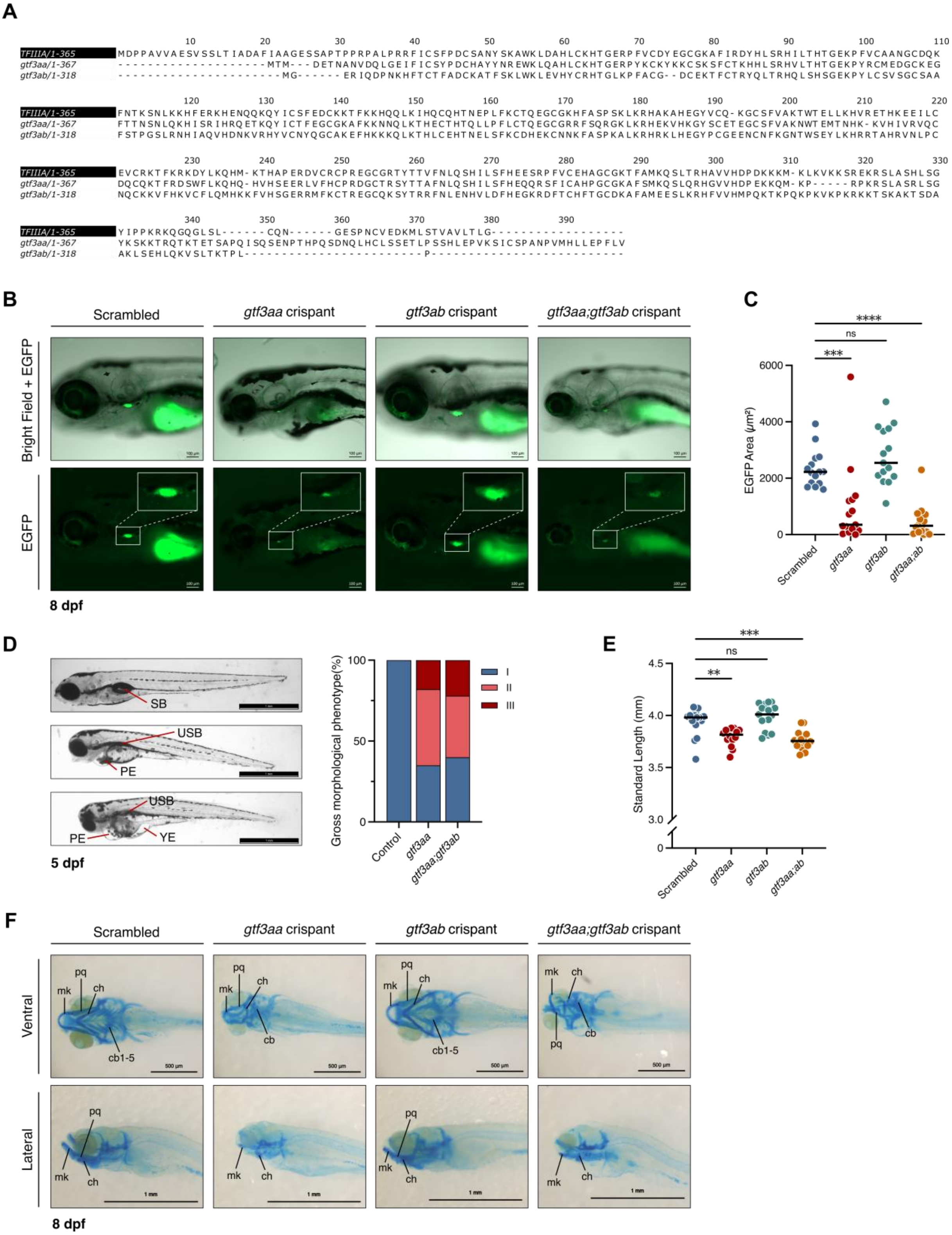


**Supplementary Figure 6 | Developmental abnormalities and early T cell developmental defects persist in *gtf3aa*-deficient zebrafish at later stages.** (**A**) Multiple sequence alignment of human TFIIIA and zebrafish ohnologs (*gtf3aa*, *gtf3ab*) generated using Clustal Omega. (**B**) Representative merged bright-field and EGFP fluorescence images, and corresponding EGFP-only images, of the thymic region in Tg(*lck*:EGFP) zebrafish larvae at 8 days post-fertilization (dpf). White boxes indicate the thymic region. (**C**) Quantification of the EGFP-positive area within the thymic region at 8 dpf. (**D**) Scoring of the gross morphological phenotype of *scrambled* control, *gtf3aa*, and *gtf3aa;gtf3ab* larvae at 5 dpf, with: (I) Normal development: Swim bladder (SB) fully inflated, no visible edema; (II) Moderate abnormalities: Swim bladder uninflated (USB) with visible pericardial edema (PE); (III) Severe abnormalities: USB with both PE and yolk sac edema (YE). (**E**) Standard body length of larvae at 8 dpf. (**F**) Representative images of lateral and ventral views of alcian blue-stained *scrambled* control, *gtf3aa*, and *gtf3aa;gtf3ab* Tg(*lck*:EGFP) larvae 8 dpf. Structures are annotated as: mk: Meckel's cartilage, pq: Palatoquadrate, ch: Ceratohyal, cb: Ceratobranchial. Data shown in (B to F) are representative of at least two independent experiments. Mean ± SD of minimum 15 fish is shown. Statistical tests: One-way ANOVA with Dunnett's multiple comparisons (C) and Kruskall-Wallis test with Dunn's multiple comparisons (E). ns: not significant, *P<0.05, **P<0.01, ***P<0.001, ****p<0.0001.

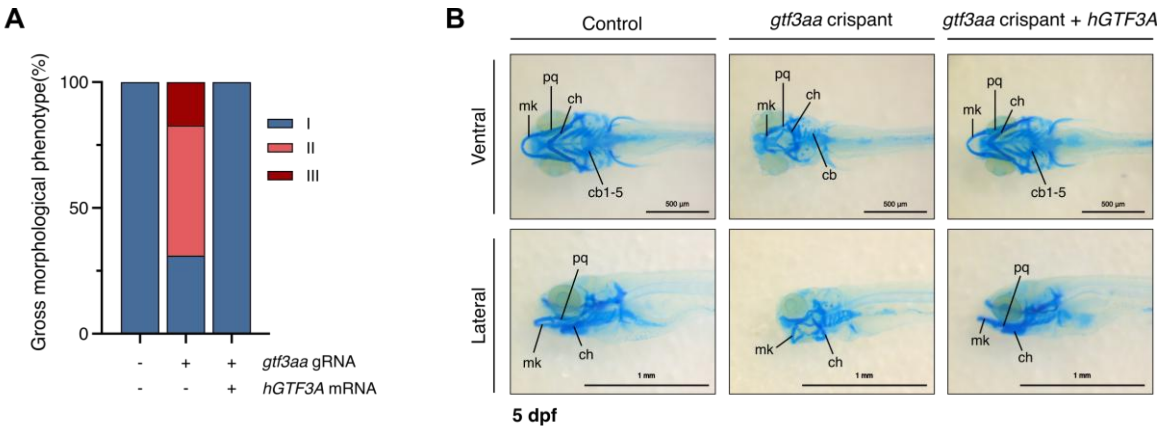


**Supplementary Figure 7 | Developmental abnormalities in *gtf3aa*-deficient zebrafish are rescued upon human *GTF3A* mRNA complementation. (A)** Developmental assessment of non-injected control larvae, *gtf3aa* crispants, and *gtf3aa* crispants co-injected with wild-type human *GTF3A* (*hGTF3A*) mRNA at 5 days post-fertilization (dpf). Larvae were categorized as: (I) Normal development: Swim bladder fully inflated, no edema; (II) Moderate abnormalities: Swim bladder uninflated with visible pericardial edema; (III) Severe abnormalities: uninflated swim bladder with both pericardial edema and yolk sac edema. **(B)** Craniofacial cartilage structures visualized by Alcian blue staining in non-injected control larvae, *gtf3aa* crispants, and *gtf3aa* crispants complemented with *hGTF3A* mRNA at 5 dpf, shown in ventral and lateral views. The right panels illustrate restoration of normal cartilage structures following *hGTF3A* complementation. Cartilage structures are annotated as: mk: Meckel's cartilage; pq: Palatoquadrate; ch: Ceratohyal; cb: Ceratobranchial. Data are representative of at least two independent experiments.

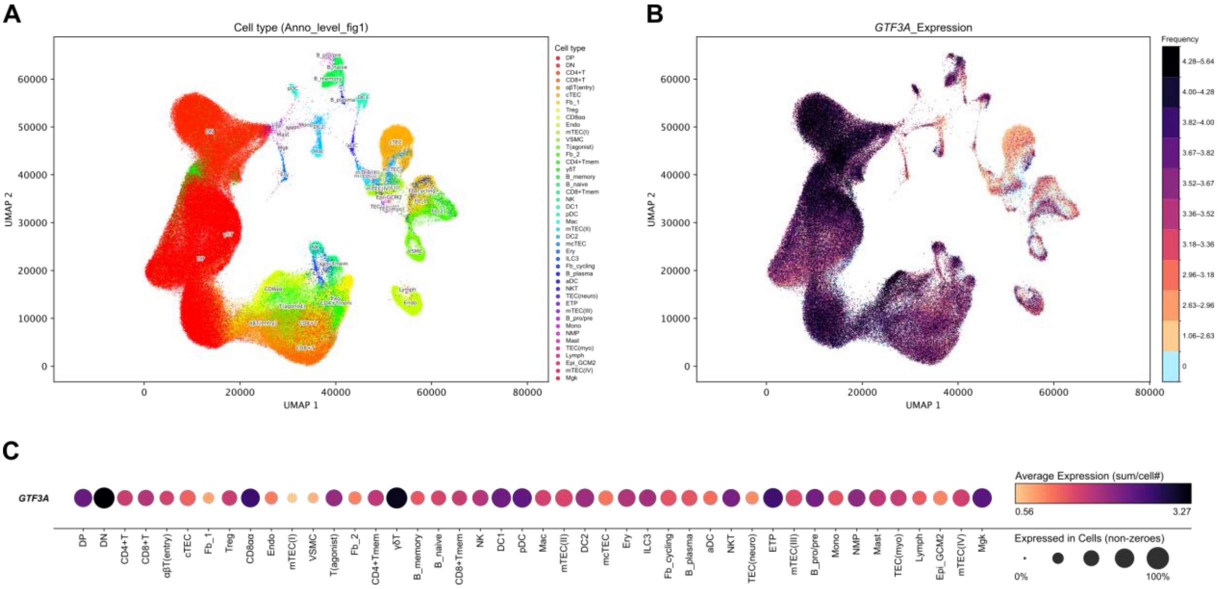


**Supplementary Figure 8 | *GTF3A* expression across cell types of the developing human thymus.** Single-cell transcriptomic data from the human thymus cell atlas (Park *et al.*, 2020), comprising 255,901 cells, were accessed through the UCSC Cell Browser (Speir *et al.*, 2021) and displayed on the atlas UMAP embedding with the published cell-type annotation (Anno_level_fig1; 44 populations). (**A**) UMAP embedding colored by cell type, with populations labeled in place. (**B**) The same UMAP colored by *GTF3A* expression using the UCSC Cell Browser *GTF3A* quantile-bin color scale. (**C**) Dot plot summarizing *GTF3A* expression by cell type. Dot size indicates the percentage of cells with non-zero *GTF3A* expression, and dot color indicates average *GTF3A* expression per cell type.

### SUPPLEMENTARY TABLES

#### Supplementary Table 1 | Treatment overview of ten TFIIIA-deficient patients from eight unrelated families.

| Patient no. | P1 | P2 | P3 | P4 | P5 | P6 | P7 | P8 | P9 | P10 |
|---|---|---|---|---|---|---|---|---|---|---|
| **Diagnosis** | **SCID** (T-B+) | **SCID** (T-B+) | **SCID** (T-B+) | **SCID** (T-B+) | **CID** | **CID** | **CID** | **CID** | **CID** | **CID** |
| **Pedigree** | Family 1 | Family 1 | Family 2 | Family 3 | Family 4 | Family 5 | Family 6 | Family 6 | Family 7 | Family 8 |
| **Treatment** | | | | | | | | | | |
| Ig substitution (IgRT) | + | + | + | + | + | + | + | + | + | - |
| Antibiotic prophylaxis | + | + | + | + | + | + | + | + | + | + |
| Fungal Prophylaxis | + | + | + | + | + | + | - | - | + | - |
| Other | - | - | Cidofovir, Ganciclovir, Maribavir | - | Growth hormone substitution (no effect), Sulfasalazine, Azathioprine | - | - | - | - | - |
| **Stem cell transplantation** | - | + | + | + | + | - | - | - | + | - |
| Age at HSCT | *NA* | 0-5y | 0-5y | 0-5y | 26-30y | *NA* | *NA* | *NA* | 6-10y | *NA* |
| Donor | *NA* | Haploidentical (mother) | MUD | Haploidentical (mother) | MUD (PBSC) | *NA* | *NA* | *NA* | MRD (sister) | *NA* |
| Conditioning regimen | *NA* | + | Alemtuzuma, treosulfan, fludarabine | Busulfan, cyclophosphamide, fludarabine | Alemtuzumab, busulfan, cyclophosphamide | *NA* | *NA* | *NA* | Fludarabine, busulfan | *NA* |
| **Outcome** | Died due to respiratory failure | Died post-transplant | Full donor chimerism at 6 months post-transplant, no GvHD | Liver VOD/MOF, Grade 4 GvHD<br>Died post-transplant | Mucositis, Pseudomonas bacteremia, squamous cell carcinoma of the tongue, norovirus diarrhea | | Mother of twins<br>Lost-to-follow up | Died due to MOF (cultures positive for Stenotrophomonas, Pseudomonas and Adenovirus) | RSV, EBV nuclear antigen IgG positive, Rhinovirus, Enterovirus<br>Skin GvHD after HSCT | Lost-to-follow up |

*EBV: Epstein-Barr virus; GvHD: Graft-versus-Host Disease; Ig: Immunoglobulin; IgRT: Immunoglobulin replacement treatment; MUD: Matched Unrelated Donor; MOF: Multiple Organ Failure; MRD: Matched Related Donor; NA: Not applicable; PBSC: Peripheral Blood Stem Cells; RSV: Respiratory syncytial virus; UnK: Unknown; VOD: Veno-Occlusive Disease*

**Supplementary Table 2 | Overview of identified *GTF3A* variants, including CADD scores, REVEL scores, and AlphaMissense predictions.**

| HGVS nomenclature (NM_002097.3) | Protein consequence | Genomic coordinates (GRCh38/hg38) | Allele frequency (gnomAD v4.1.0) | CADD score | REVEL score | AlphaMissense | ClinVar (Accession) |
|---|---|---|---|---|---|---|---|
| c.190C>G | p.H64D | *Not present* | *Private* | 31.0 | 0.600 | 0.9891 | Not reported |
| c.322G>T | p.D108Y | 13-27429889-G-T | 0.000008491 | 25.5 | 0.215 | 0.2335 | Not reported |
| c.358C>T | p.H120Y | 13-27429925-C-T | 0.00000129992 | 25.3 | 0.595 | 0.9329 | Not reported |
| c.435del | p.K145Nfs*5 | 13-27430565-GA-G | 0.00000310116 | 32.0 | − | − | Not reported |
| c.541C>T | p.R181* | 13-27432783-C-T | 0.000006231 | 38.0 | − | − | Not reported |
| c.585T>G | p.C195W | *Not present* | *Private* | 27.5 | 0.559 | 0.9592 | Not reported |
| c.655T>C | p.C219R | 13-27434816-T-C | 0.00000747166 | 28.8 | 0.909 | 0.9894 | Not reported |
| c.682_683dup | p.K229Afs*9 | *Not present* | *Private* | 34.0 | − | − | Not reported |
| c.722dup | p.E242Rfs*21 | 13-27434879-G-GC | 0.00012584 | 26.2 | − | − | Not reported |
| c.814_816del | p.E272del | *Not present* | *Private* | 20.1 | − | − | Not reported |
| c.956G>C | p.R319P | 13-27435455-G-C | 0.00000062 | 32.0 | 0.345 | 0.4209 | Not reported |
| c.956G>A | p.R319Q | 13-27435455-G-A | 0.000218275 | 32.0 | 0.164 | 0.1851 | Not reported |
| c.955C>T | p.R319W | 13-27435454-C-T | 0.000006201 | 28.0 | 0.155 | 0.2983 | Uncertain significance (VCV002531718.2) |
| c.1090del | p.G365Afs*64 | 13-27435586-AC-A | 0.00000371806 | 22.4 | − | − | Not reported |

#### Supplementary Table 3 | Overview of homozygous *GTF3A* variants in gnomAD v4.1.0.

| Genomic coordinates (GRCh38/hg38) | rsID (dbSNP) | HGVS nomenclature (NM_002097.3) | Protein consequence | Allele frequency (gnomAD v4.1.0) | Number of homozygotes (gnomAD v4.1.0) | CADD score | ClinVar (Accession) |
|---|---|---|---|---|---|---|---|
| 13-27434894-G-C | rs7323 | c.733G>C | p.V245L | 0.253521836 | 54596 | 21.6 | Not reported |
| 13-27429881-A-C | rs1803810 | c.314A>C | p.N105T | 0.005685849 | 448 | 6.15 | Not reported |
| 13-27429961-T-C | rs188074983 | c.394T>C | p.Y132H | 0.003131099 | 9 | 24.8 | Not reported |
| 13-27434801-C-T | rs189742565 | c.644-4C>T | − | 0.001372792 | 8 | 1.79 | Not reported |
| 13-27427168-C-T | rs201449609 | c.278C>T | p.T93I | 0.000950473 | 4 | 2.12 | Not reported |
| 13-27434805-A-G | rs200137415 | c.644A>G | p.E215G | 0.001013158 | 4 | 23.5 | Not reported |
| 13-27427155-C-T | rs551663231 | c.265C>T | p.R89C | 0.00017505 | 3 | 23.5 | Not reported |
| 13-27427161-A-C | rs766824645 | c.271A>C | p.I91L | 0.00000312 | 1 | 0.054 | Not reported |
| 13-27435130-T-G | rs767973928 | c.874-3T>G | − | 0.0000385 | 1 | 17.6 | Not reported |
| 13-27435523-T-G | rs200106325 | c.1024T>G | p.C342G | 0.0000818 | 1 | 13.1 | Not reported |

**Supplementary Table 4 | Sequences of primers used for reverse-transcription quantitative PCR (RT-qPCR) analysis.**

| Primer sequence (5' → 3') | Reference | Anneal site | Comment |
|---|---|---|---|
| ***Reverse-transcription qPCR analysis (RT-qPCR)*** | | | |
| Forward: GGAGAAAAGCCGTTTGTTTG<br>Reverse: TGGGTGATGCAAAGTGTTTC | NM_002097.3 (*GTF3A*) | Exon 2<br>Exon 5 | Primer pair |
| Forward: TACGGCCATACCACCCTGAA<br>Reverse: GCGGTCTCCCATCCAAGTAC | NR_023363.1 (*5S rRNA*) | -<br>- | Primer pair |
| ***Housekeeping genes RT-qPCR*** | | | |
| Forward: CCAACCGCGAGAAGATGA<br>Reverse: CCAGAGGCGTACAGGGATAG | NM_001101.5 (*ACTB*) | - | Primer pair |
| Forward: CACCCACTCCTCCACCTTTGA<br>Reverse: GTCCACCACCCTGTTGCTGTAG | NM_002046.7 (*GAPDH*) | - | Primer pair |
| Forward: CCTGGCGTCGTGATTAGTGA<br>Reverse: TCTCGAGCAAGACGTTCAGT | NM_000194.3 (*HPRT*) | - | Primer pair |

**Supplementary Table 5 | Fluidigm antibodies for deep immunophenotyping using CyTOF.**

| Antigen | Tag | Clone | Company | Catalog number |
|---|---|---|---|---|
| CXCR3 | 163Dy | G025H7 | Fluidigm | 3163004B |
| TCRγδ | 152Sm | 11F2 | Fluidigm | 3152008B |
| CD19 | 142Nd | HIB19 | Fluidigm | 3142001B |
| CD38 | 144Nd | HIT2 | Fluidigm | 3144014B |
| CD123 | 151Eu | 6H6 | Fluidigm | 3151001B |
| Vα7.2 | 153Eu | 3C10 | Fluidigm | 3153024B |
| CD3 | 154Sm | UCHT1 | Fluidigm | 3154003B |
| CD45RA | 155Gd | HI100 | Fluidigm | 3155011B |
| CD27 | 158Gd | L128 | Fluidigm | 3158010B |
| CD1c | 159Tb | L161 | Biolegend | 331502 |
| CLEC9A | 161Dy | 8F9 | Fluidigm | 3161018B |
| CD161 | 164Dy | HP-3G10 | Fluidigm | 3164009B |
| CD8 | 168Er | SK1 | Fluidigm | 3168002B |
| iNKT | 170Er | 6B11 | Fluidigm | 3170015B |
| CCR4 | 175Lu | L291H4 | Fluidigm | 3175035A |
| CD4 | 174Yb | SK3 | Fluidigm | 3174004B |
| CD21 | 162Dy | REA940 | Miltenyi Biotec Inc. | 130-124-315 |
| NKG2C | 165Ho | REA205 | Miltenyi Biotec Inc. | 130-122-278 |
| CD20 | 148Nd | 2H7 | Biolegend | 302302 |
| HLA-DR | 173Yb | L243 | Fluidigm | 3173005B |
| CCR10 | 156Gd | REA326 | Miltenyi Biotec Inc. | 130-122-317 |
| CD45 | 089Y | HI30 | Fluidigm | 3089003B |
| CD66b | 116Cd | QA17A51 | Biolegend | 396902 |
| CCR6 | 141Pr | G034E3 | Fluidigm | 3141003A |
| CD127 | 143Nd | A019D5 | Fluidigm | 3143012B |
| CD11c | 147Sm | Bu15 | Fluidigm | 3147008B |
| CD25 | 149Sm | 2A3 | Fluidigm | 3149010B |
| NKVFS1 | 150Nd | NKVFS1 | Bio Rad | MCA2243GA |
| CCR7 | 167Er | G043H7 | Fluidigm | 3167009A |
| NKG2A | 169Tm | Z199 | Fluidigm | 3169013B |
| CXCR5 | 171Yb | RF8B2 | Fluidigm | 3171014B |
| CD24 | 166Er | ML5 | Fluidigm | 3166007B |
| CD31 | 145Nd | WM59 | Fluidigm | 3145004B |
| CD14 | 160Gd | M5E2 | Fluidigm | 3160001B |
| CD56 | 176Yb | NCAM16.2 | Fluidigm | 3176008B |
| CD57 | 172Yb | HNK-1 | Biolegend | 359602 |
| KIR3DL1L2 | 150Nd | REA970 | Miltenyi Biotec Inc. | 130-126-489 |
| IgD | 146Nd | IA6-2 | Fluidigm | 3146005B |
| CD16 | 209Bi | 3G8 | Fluidigm | 3209002B |

**Supplementary Table 6 | Detailed overview of the monoclonal antibodies used for immunophenotyping.**

| Antigen | Fluorochrome | Clone | Company | Catalog number |
|---|---|---|---|---|
| CD24 | BUV395 | ML5 | BD Biosciences | 563818 |
| CD8 | BUV496 | RPA-T8 | BD Biosciences | 612942 |
| CD19 | BUV496 | SJ25C1 | BD Biosciences | 612939 |
| CCR7 | BUV615 | 2-L1-A | BD Biosciences | 751099 |
| HLA-DR | BUV661 | G46-6 | BD Biosciences | 612981 |
| CD95 | BUV737 | 563 | BD Optibuild | 741868 |
| CD4 | BUV805 | SK3 | BD Biosciences | 612887 |
| CD25 | BV421 | M-A251 | BD Biosciences | 562442 |
| CXCR5 | BV480 | RF8B2 | BD Biosciences | 566191 |
| CD16 | BV570 | 3G8 | Biolegend | 302036 |
| CD31 | BV605 | WM98 | BD Biosciences | 562855 |
| CD11c | BV650 | 3.9 | Biolegend | 301638 |
| CD3 | BV711 | UCHT1 | Biolegend | 300464 |
| CD14 | BV750 | 63D3 | Biolegend | 367136 |
| CD161 | BV785 | HP-3G10 | Biolegend | 339930 |
| TCRγδ | FITC | B1 | Biolegend | 331208 |
| CD123 | BB630 | 7G3 | BD Horizon | 624294 |
| IgD | BB700 | IA6-2 | BD Biosciences | 566539 |
| CD56 | BB790-P | NCAM16-2 | BD Biosciences | 624296 |
| FoxP3 | PE | 206D | Biolegend | 320108 |
| CD127 | PE-Dazzle594 | A019D5 | Biolegend | 351336 |
| SA | PE-Cy5 | 206D | BD Biosciences | 554062 |
| CD27 | PE-Cy7 | O323 | Biolegend | 302838 |
| CD45RA | APC | HI100 | Biolegend | 304112 |
| Vα7.2 | AF700 | 3C10 | Biolegend | 351728 |
| CD38 | APC-Fire750 | HIT2 | Biolegend | 303546 |
| Vα24Jα18 | biotin | 6B11 | Thermofisher | 13-5806-82 |

**Supplementary Table 7 | Detailed overview of the antibodies used for immunophenotyping the T cell compartment derived from artificial thymic organoids (ATO).**

| Antigen | Fluorochrome | Clone | Company | Catalog number |
|---|---|---|---|---|
| CD56 | BV421 | 5.1H11 | Biolegend | 362551 |
| CD56 | FITC | MEM-188 | Biolegend | 304604 |
| TCRαβ | BV510 | IP26 | Biolegend | 306734 |
| TCRαβ | PerCPCy5.5 | IP26 | Biolegend | 306724 |
| CD1a | BV711 | HI149 | Biolegend | 300139 |
| CD1a | APC | HI149 | Biolegend | 300110 |
| CD8β | PE | REA715 | Miltenyi | 130-110-568 |
| CD8β | PE | 2ST8.5H7 | BD Biosciences | 641057 |
| Live/Death (Propidium iodide) | PE Texas red | - | ThermoFisher | J66764.MC |
| Live/Death | Qdot 565 | - | ThermoFisher | L34959 |
| CD45 | PerCpCy5.5 | HI30 | Biolegend | 304028 |
| CD45 | V500 | HI30 | BD Biosciences | 560779 |
| CD5 | PE-Cy7 | UCHT2 | Biolegend | 300622 |
| CD5 | PE-Cy7 | UCHT2 | ThermoFisher | 25-0059-42 |
| CD4 | APC | RPA-T4 | Biolegend | 300514 |
| CD4 | APC-Cy7 | SK3 | BD Biosciences | 341095 |
| CD7 | AF700 | M-T701 | Becton Dickinson | 561603 |
| CD7 | AF700 | eBio124-1D1 | ThermoFisher | 56-0079-42 |
| CD3 | APC-Cy7 | UCHT1 | Biolegend | 300426 |
| CD3 | BV421 | UCHT1 | BD Biosciences | 562426 |

**Supplementary Table 8 | CRISPR RNAs (crRNAs) sequences for the generation of crispants.**

| zebrafish model | crRNAs | Target site |
|---|---|---|
| *gtf3aa* crispant | AGGAGCGAGTGCACCCGTCT | Exon 7 |
| *gtf3ab* crispant | CTTGAGGAACCACATTGCTC | Exon 3 |
| *scrambled* | GCAGGCAAAGAATCCCTGCC | - |

**Supplementary Table 9 | Sequences of primers used for genotyping (Sanger sequencing) of crispants.**

| Primer sequence (5' → 3') | Reference |
|---|---|
| ***gtf3aa*** | |
| Forward: TCTCTCAGTTAAACAGAAATTGGG<br>Reverse: CATAATATCAGTCTATTGCGCCG | NM_001003866.1 |
| ***gtf3ab*** | |
| Forward: TCTGTGATGTGGTTTACTGCG<br>Reverse: TTGCACATCCTTGATAGTTGC | NM_001089544.2 |
